# PREFER-IT: A transdisciplinary co-created framework to realise inclusive medical AI

**DOI:** 10.1101/2025.11.03.25339443

**Authors:** Patrícia Pita Ferreira, Sara Soriano Longarón, Wiam Bouisaghouane, Jetse Goris, Anne H. Hoekman, Balázs Markos, Benjamin Maus, Giorgia Pozzi, Hadi Hasan, Indre Kalinauskaité, Jonáh Stunt, Joosje D. Kist, Judith van der Elst, Katell Maguet, Liv Ziegfeld, Maarten Cuypers, Megan Milota, Michelle Habets, Sara Colombo, Špela Petrič, Steff Groefsema, Steven Warmelink, Elja Daae, Giovanni Briganti, Ildikó Vajda, Matias Valdenegro-Toro, Matthias Braun, Pieter Jeekel, Simone Goosen, Alex Schepel, Laxmie Ester, Riane Kuzee, Sophie de Klerk, Claudine Lamoth, Lisa Ballard, Mirjam Plantinga

## Abstract

Artificial intelligence (AI) in healthcare holds transformative potential but risks exacerbating existing health disparities if inclusivity is not explicitly accounted for. This study addresses the disconnected discussions on inclusive medical AI by developing a comprehensive framework, PREFER-IT. This framework is based on the outcomes of a five-day transdisciplinary co-creation workshop that involved 37 experts from diverse backgrounds, including healthcare, ethics, law, social sciences, AI, and patient advocacy. For this workshop, we used design thinking and participatory methodologies to develop a framework for realising inclusive medical AI. We identified three key challenges for realising inclusive medical AI: integrating the lived experiences and stakeholder voices across the AI lifecycle, designing data collection practices that promote fairness and prevent inequalities, and fostering regulatory frameworks to uphold human rights and promote inclusivity. The analysis of participants’ perspectives informed the development of eight key thematic clusters of PREFER-IT: Participatory and co-design approaches (P), Representative and diverse data (R), Education and digital literacy (E), Fairness (F), Ethical and legal accountability (E), Real-world validation and feedback (R), Inclusive communication (I), and Technical interoperability (T). These elements were mapped across structural layers of AI (humans, data, system, process, and governance) and the AI lifecycle to guide inclusive design, development, validation, implementation, monitoring, and governance. This framework fosters stakeholder engagement and systemic change, positioning inclusion as a guiding principle in practice. PREFER-IT offers a practical and conceptual contribution for how to include ethical, legal and societal aspects when aiming to foster responsible and inclusive AI in healthcare.

**Author Summary:** Artificial intelligence (AI) is being used more and more in healthcare to improve diagnosis, treatment, and personalised care. However, if not designed carefully, these technologies can unintentionally increase existing inequalities and exclude certain groups from their benefits. In our study, we brought together experts from healthcare, ethics, law, social sciences, and patient advocacy to explore how AI in medicine can be made more inclusive. Over five days, we worked together to identify key issues and come up with practical solutions.

We focused on three main areas: 1) Ensuring diverse voices are heard during the development of AI tools; 2) Making data collection fair and representative; and 3) Creating regulations that protect human rights.

From the discussions of the workshop, we created the PREFER-IT framework, which outlines eight key principles for inclusive AI:

- **P**articipatory and co-design approaches
- **R**epresentative and diverse data
- **E**ducation and digital literacy
- **F**airness
- **E**thical and legal accountability
- **R**eal-world validation and feedback
- **I**nclusive communication
- **T**echnical interoperability

This framework helps guide developers, policymakers, and healthcare professionals in creating AI systems that are not only effective but also fair and respectful of all users. Our work emphasises the importance of involving patients and communities in shaping the future of AI.

## Introduction

The advances and applications of artificial intelligence (AI) in healthcare, referred to in this paper as *medical AI*, have the potential to transform society. Medical AI could improve diagnosis, personalise treatment, increase operational efficiency, optimise resources such as staff time, equipment, and hospital capacity, and improve patients’ experience [1–5]. Despite these possible benefits, medical AI carries known risks: it may reinforce existing biases against historically disadvantaged populations, underserved groups, and individuals experiencing (multiple) layers of vulnerability [6], and it remains prone to errors and patient harm [7–9]. If left unaddressed, these risks could ultimately exacerbate health disparities. [1]. Furthermore, medical AI raises questions regarding accessibility, autonomy, equity, human rights, and inclusivity [1]. Inclusivity is a recognised principle in healthcare, but concerns remain that AI may reproduce or exacerbate disparities across patient and population groups [7,10]. Groups such as older adults, ethnic minorities, and individuals with disabilities may face disproportionate risks or limited benefits from AI innovations [1,10–12].

The literature shows a growing attention to issues of bias, trust, and fairness in AI [2,3,8,9,11,13–22], with different disciplines addressing these issues from complementary perspectives. Medical informatics research prioritises algorithmic fairness metrics [7,9] and technical bias mitigation [16,23,24], focusing on the iterative phases of the AI lifecycle, which encompass the design, development, validation, implementation, and monitoring of AI [2], while often not addressing social determinants of health and inequity. Ethics, legal, and social science literature emphasise autonomy, justice, and human rights, frequently lacking operationalisation into technical or clinical practice [25–27]. Clinical and public health research addresses data representativeness, patient outcomes, and health disparities, often concentrated in high-income contexts [7,10,26].

While each field contributes valuable insights to these issues, these contributions have largely evolved in parallel, resulting in a fragmented landscape of efforts that, while individually valuable, collectively limit the development of cohesive and actionable solutions. This fragmentation results in uneven attention to people experiencing (multiple) layers of vulnerabilities [6] (e.g., more emphasis on race and gender bias than on age, disability or their intersections [12,28]) and gaps across the AI lifecycle, with deployment, monitoring, and governance being less explored than data collection and model design, for example [26]. Moreover, current approaches rarely engage with broader notions of inclusion, such as the social and structural contexts in which AI tools operate [12,29] or the systemic, participatory integration of marginalised voices and needs across the AI lifecycle.

To build on these diverse contributions, this paper adopts the lens of *inclusive medical AI* to examine how principles of inclusion can be systematically embedded into the AI lifecycle and governance of AI in healthcare.

We define medical inclusivity as the equal ability, opportunity and right of all individuals, particularly those who are underserved, vulnerable or otherwise disadvantaged based on identity and/or circumstances, to access health services, receive compassionate and high-quality care, and achieve equitable health outcomes, in ways that respect human dignity. [13]. As a prerequisite for medical inclusivity, AI systems must be designed and implemented to ensure equitable access and use across all population groups, regardless of attributes such as age, sex, gender, income, race, ethnicity, sexual orientation, disability, or other characteristics protected by human rights principles [1].

Some works, although not specifically focused on healthcare, have attempted to address this gap by exploring diversity and inclusion by design [9,24,25] or by advocating for stakeholder engagement in design and evaluation [13,30]. For example, Zowghi and da Rimini propose five pillars (humans, data, process, system, governance) for embedding diversity and inclusion by design across AI ecosystems. This approach extends beyond fairness metrics by urging the inclusion of diverse voices, contexts, and institutions [30]. Wang and Blok propose a multi-level framework that shifts from AI micro-level issues (e.g., dataset bias, algorithm transparency) to structural concerns, from meso (clinical and organisational) to macro (systemic and socio-political) levels, which shape AI’s broader impacts [25].

The FUTURE-AI guideline represents a first step, offering a structured, global consensus on trustworthy AI in healthcare across the AI lifecycle. While it addresses fairness, universality, and stakeholder engagement, its implementation of inclusivity remains limited: inclusivity is mainly viewed through bias mitigation and technical fairness metrics, rather than as a systematic design principle rooted in different contexts, lived experiences, and societal perspectives. [2]

For healthcare specifically, the field remains fragmented, where valuable yet dispersed insights still need to be synthesised into a comprehensive, pragmatic framework for inclusive medical AI. While technical and governance-oriented guidelines exist [2], and ethical principles have been articulated in policy documents [1,11], systematic approaches to realise inclusivity remain dispersed and underdeveloped. General frameworks on inclusivity are not always transferable to healthcare, given the high-stakes and sensitive nature of this domain [27,32]. Medical AI tools have a direct impact on health outcomes, and the risks are substantial. Furthermore, healthcare is characterised by hierarchies and power dynamics, knowledge asymmetries between experts and patients, and the influence of professional and institutional norms [33–35]. These conditions can lead to the exclusion of voices and lived experiences, without explicit mechanisms for medical inclusivity.

This paper bridges this gap by presenting a set of co-created foundational requirements, named **PREFER-IT**, for the design, development, validation, implementation, monitoring, and governance of inclusive medical AI. Developed through a transdisciplinary co-creation process involving multiple disciplines and stakeholder perspectives, including patients, PREFER-IT provides a practical framework to realise medical AI inclusivity. In this paper, we describe how PREFER-IT was developed.

## Methods

### Recruitment of participants

To explore the challenges of inclusive medical AI and identify the requirements to address them, we organised a five-day transdisciplinary workshop, held from 10 to 14 February 2025 at the Lorentz Centre in Leiden (The Netherlands). The event brought together 37 experts, of whom six were organisers/facilitators, eight were (online) motivational speakers, and three were patient experts. The areas of expertise included the following categories: 1 – healthcare and public health; 2 – ethics, law and policy; 3 – social sciences, psychology, humanities and design; 4 – AI and data science; 5 – patient experts and advocacy. We employed a combination of purposive and snowball sampling to recruit participants, and experts were selected for their expertise in responsible AI and/or inclusive health(care). Attention was paid to ensuring diversity in disciplinary background, area of specialisation, seniority, geographical location, and gender, to foster multi-perspective dialogue (S1 Fig). The second day of the workshop included on-site patient experts from different underserved groups.

### Preparation and priority setting of the workshop

We used the design thinking methodology, adapting the double diamond model and the innovation and systemic design frameworks (Fig 1). Central to this method is the iterative phases for problem exploration and solution development, with the inclusion of all stakeholders [36–39]. The design thinking method was complemented by introducing a systemic approach for realising inclusive medical AI, based on the different levels (micro, meso, macro), as proposed by Wang and Blok [25]. Additionally, the workshop aimed to integrate scientific insights with patient perspectives to promote transdisciplinary discussions, deepen understanding of the problem, and generate potential solutions or requirements (also known as ideation) for inclusive medical AI. This structured approach facilitated knowledge exchange and collaboration across disciplines.

**Fig 1.**
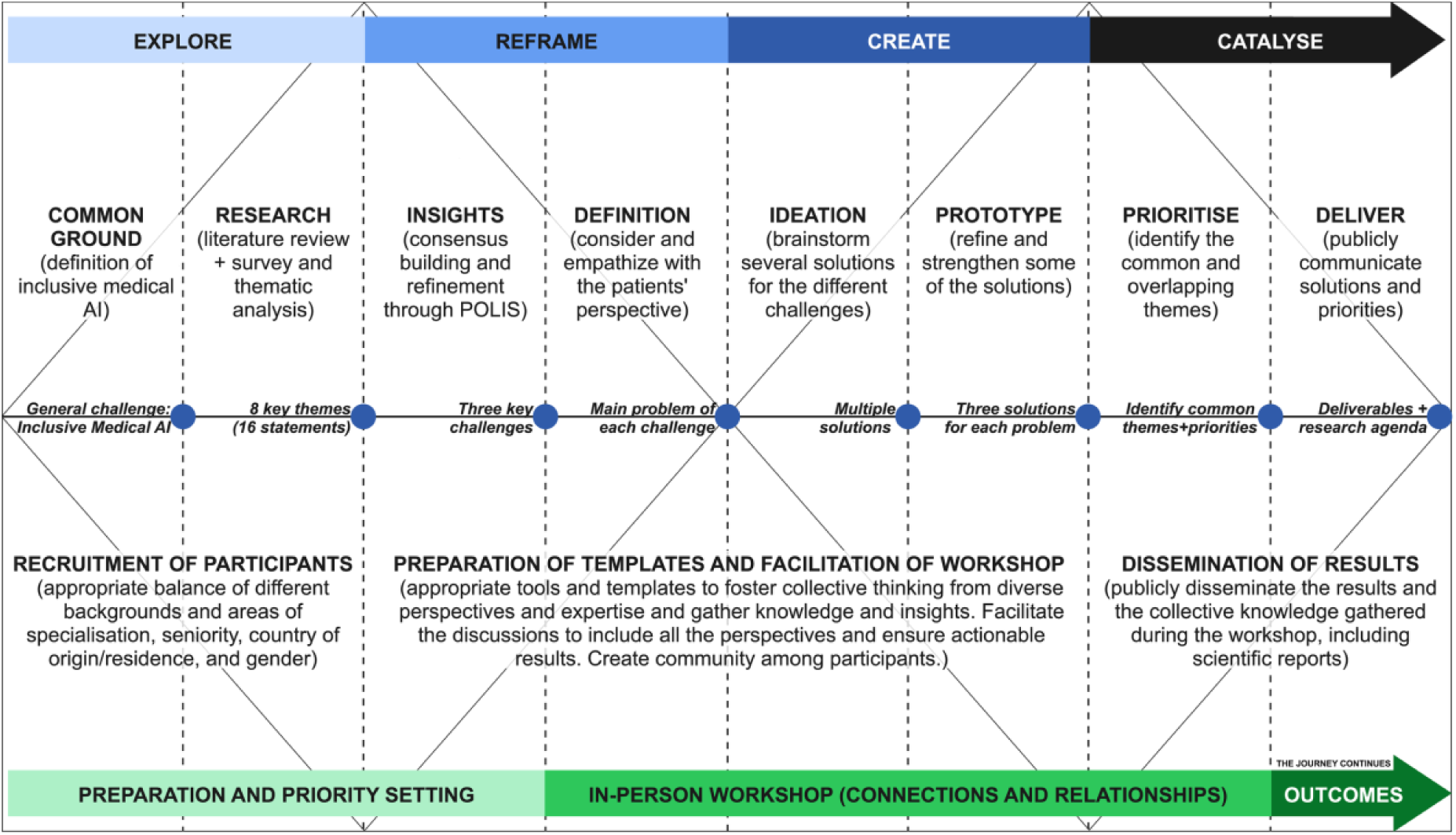
Structured workflow for the workshop. Adaptation of the double diamond model and the innovation and systemic design frameworks

Fig 1 describes the framework for the different stages of the workshop, based on the design thinking methodology [36–38]. S2 Fig presents the adapted Wang and Blok framework, showing how the identified challenges of inclusive medical AI and their proposed requirements relate across multiple levels [40].

In the preparation and priority-setting stages, participants completed a short anonymous online survey with three open-ended questions: how they understood inclusive medical AI, what they considered necessary to realise it, and which topics should be prioritised in the workshop.

The first question informed our working definition of medical inclusivity, which was presented in the introduction section (*Common Ground* stage). The second and third questions focused on the perceived needs and challenges associated with realising inclusive medical AI. We conducted a thematic analysis of participants’ responses, which led to the identification of eight themes related to challenges for realising inclusive medical AI (*Research* stage).

To focus the discussion into a set of sub-challenges that could be effectively addressed during the in-person workshop, we performed a consensus-building exercise. For this, we started by drafting 16 statements (two per theme) from the thematic analysis, supported by ChatGPT-4o (OpenAI) for coding and phrasing. We then refined these statements through iterative cross-checking with the original survey data and the expertise of the research team. The resulting statements captured individual ideas related to the challenges of inclusive medical AI and were subsequently used in Pol.is, an open-source digital platform for consensus building [41]. Pol.is collects and analyses opinions using machine learning, applying dimension reduction and clustering to identify average views and detect distinct opinion groups [41]. Participants could also contribute with their own statements, resulting in seven new anonymous submissions, all aligned with the original themes. A complete list of the statements and their corresponding themes is provided in Table 1.

**Table 1.**
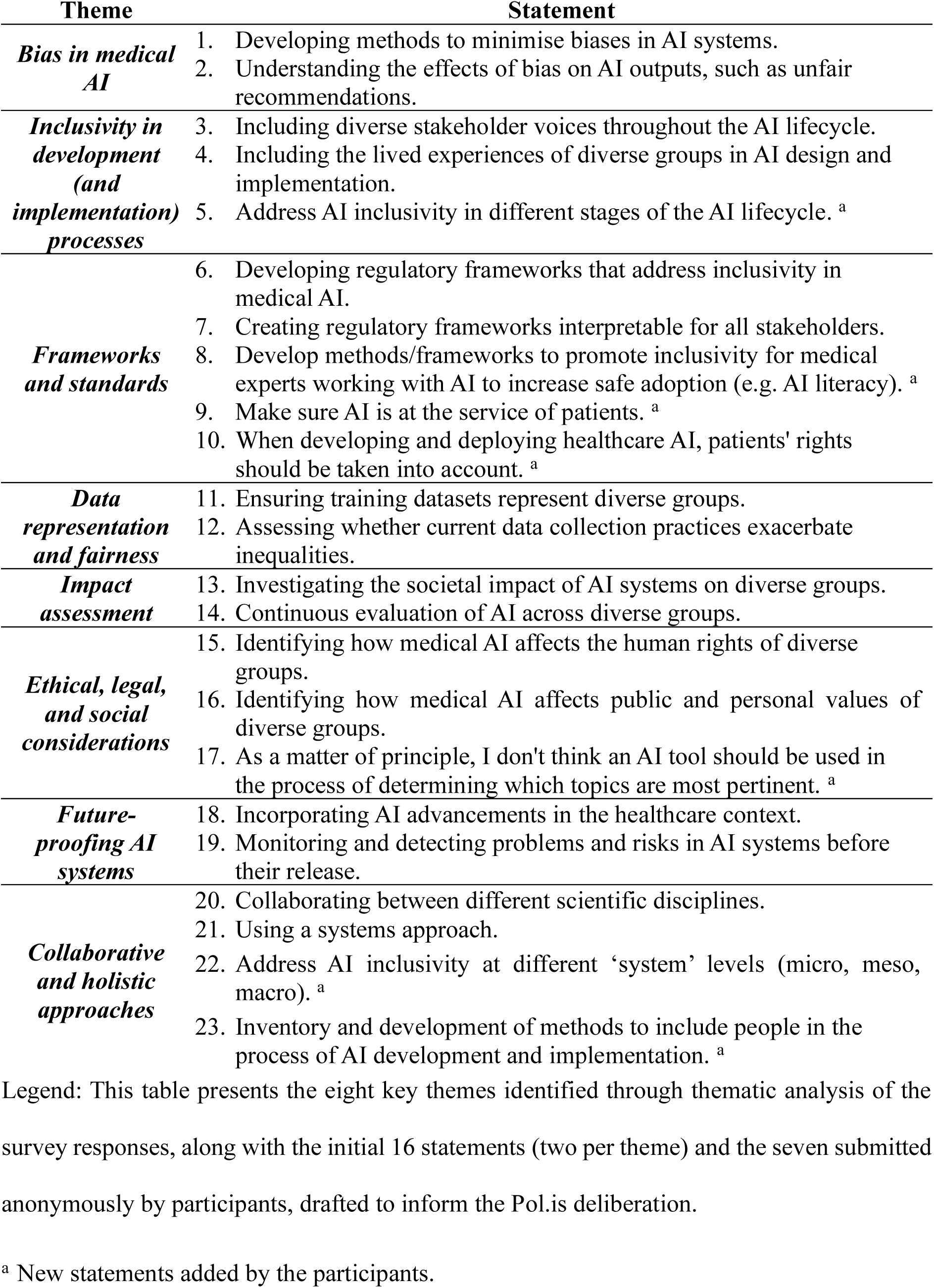
Themes and statements on inclusive medical AI used in Pol.is.

To include patient voices in the preparation and priority-setting stages, a focus group and a separate consensus-building exercise were conducted with a Patient and Public Involvement (PPI) group. During the consensus-building phase, participants used the Pol.is platform to share their perspectives and priorities in lay language. Their preferences were incorporated into the final set of sub-challenges on inclusive medical AI prioritised for the workshop.

Participants of the workshop were asked to vote, through Pol.is, on each statement based on what they considered the most pressing challenges to address during the five-day in-person workshop (*Insights* stage).

Based on the consensus-building exercise through Pol.is (full results in S1 Table), we refined the sub-challenges considered a priority for the workshop. In addition to considering the level of agreement among participants and the feasibility of addressing the sub-challenges within the five-day format, we applied a set of criteria established after reviewing the Pol.is results. These criteria ensured that each selected sub-challenge was:

1. Distinctive, addressing unique and complementary aspects of the workshop’s overarching goal;
2. Balanced between human and technical dimensions, grounded in patient needs and lived experiences;
3. Interdisciplinary, suitable for engagement across diverse fields of expertise;
4. Aligned with the working definition of inclusive medical AI, reflecting the core pillars of ability, opportunity, and dignity for equitable health outcomes;
5. Actionable and impactful, enabling actionable insights and identifying research gaps; and
6. Reflective of the PPI perspective, consistent with priorities identified by the PPI focus group.

Three challenges with each a corresponding research question were then identified to guide the work and discussions of the participants during the in-person workshop (*Insights* stage):

A. **Integrating lived experiences and stakeholder voices across the AI lifecycle:** *How can we ensure that the lived experiences of diverse groups and stakeholder voices are systematically included throughout the AI lifecycle?*
B. **Designing data collection practices to promote fairness and prevent inequalities:** *How can we ensure that data collection practices are representative, fair, and equitable and do not exacerbate inequalities while minimising biases in AI systems?*
C. **Fostering regulatory frameworks to uphold human rights and promote inclusivity:** *How can we ensure that regulatory and ethical frameworks are used in AI development and implementation to safeguard human rights, address ethical concerns, promote trust, and support inclusive practices across the AI lifecycle?*

### In-person workshop

The participants were divided into three groups during the in-person workshop, ensuring a balanced distribution of expertise. Each group was asked to collaborate in developing potential requirements and solutions for each of the three challenges related to inclusive medical AI. The work of each group was guided by facilitators, using templates based on design thinking, which are summarised in S3 Fig.

Data was collected on sticky notes by the participants or by the facilitators. Each day, the collected data from each group was uploaded as photographs to an online research drive and directly to a Miro board [42], ensuring that all participants could follow the group’s progress. *During day one*, participants engaged in a structured, collaborative process to refine the three predefined challenges (*Definition* stage). Through facilitated brainstorming, each group deconstructed their assigned challenge into sub-problems, clustered them thematically, and mapped interconnections. To prioritise the most salient sub-problems, we used a consensus-based approach, complemented by dot-voting when needed (a participatory technique where participants visually cast votes with stickers to indicate their preferred options). Based on these, groups formulated preliminary problem statements, drawing on their disciplinary expertise and professional experience. Stakeholder mapping was conducted to identify key actors and relationships relevant to each challenge.

*On the second day,* three patient experts joined the workshop to co-create with participants. Each group collaborated with a patient to develop a patient persona and journey map, refining the initial problem statements from a patient-centred perspective. Personas (fictional yet evidence-informed representations of patient types) were used to capture diverse needs and experiences, while patient journey maps illustrated the steps and challenges individuals face within healthcare systems. This process enabled the group to explore how inclusive medical AI could better support patients across different circumstances.

*The third day* was dedicated to the *Ideation* stage. Participants initially generated ideas, through silent ideation, for solving the challenges individually, then participated in group-based clustering, elaboration, and synthesis. They were encouraged to contribute ideas across groups, facilitating cross-pollination between challenges and enhancing transdisciplinary dialogue.

*On the fourth day,* participants focused on developing solutions (*Prototype* stage). Each group selected a subset of ideas generated during the ideation phase and elaborated on at least three in greater detail. This involved defining minimum implementation conditions, including roadmaps, technical and organisational requirements, enabling features, anticipated challenges, performance metrics, cost considerations, and stakeholder engagement strategies. The resulting prototypes were pitched to the whole group, allowing for iterative refinement based on collective feedback.

*The final day* focused on identifying common themes among the developed solutions and establishing shared priorities for advancing inclusive medical AI through consensus (*Prioritise* stage). Participants collaboratively synthesised overlapping themes and reflected on missing elements. This concluding session consolidated workshop outcomes, with a strong emphasis on sustained patient involvement and the realisation of inclusivity principles throughout the AI lifecycle.

*After the workshop*, we analysed the raw data. The most important findings and the collective insights from the workshop are presented in the results section (*Deliver* stage).

### Declaration of Generative AI and AI-assisted technologies

During the workshop preparation, the authors used ChatGPT-4o (OpenAI) to help conduct a preliminary thematic analysis of the survey sent to the participants and draft the preliminary initial statements fed into Pol.is. ChatGPT-4o (OpenAI) was also used to simplify and annotate the code to analyse raw data and generate S1 Table and S1 Fig in R statistical software (version 4.4.3; R Foundation for Statistical Computing). While preparing this manuscript, the authors used AI-assisted tools (e.g Grammarly) to check grammar and spelling and improve readability. After using these tools, the authors reviewed and edited the content as needed and take full responsibility for the content of the published article.

## Results

The workshop and the development of the PREFER-IT framework followed an iterative co-creation process involving the stages of *definition*, *ideation*, *prototype*, and *prioritisation*. This process culminated in a final phase of the *PREFER-IT framework consolidation* based on the thematic analysis and clustering of the workshop data. The collective insights and results from the in-person workshop are summarised in Fig 2.

**Fig 2.**
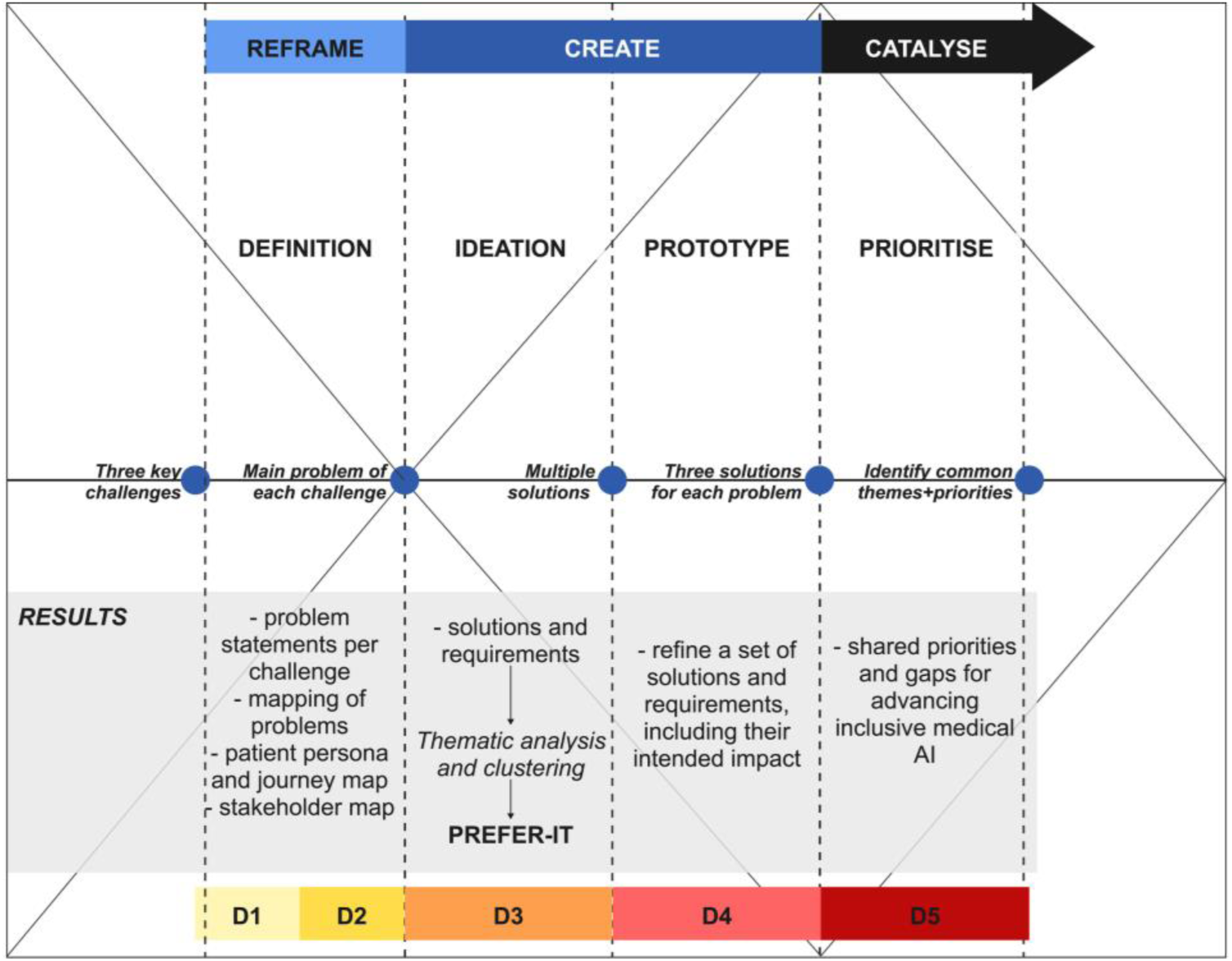
Results of the workshop summarised by day (D). The collective insights and main outcomes of the in-person workshop, by day.

### Definition

Table 2 outlines the problem clustering, the problem statements and the stakeholder map for each challenge. Across all three predefined challenges, participants identified a set of converging concerns regarding the inclusivity of AI in healthcare (*Definition* stage).

**Table 2.**
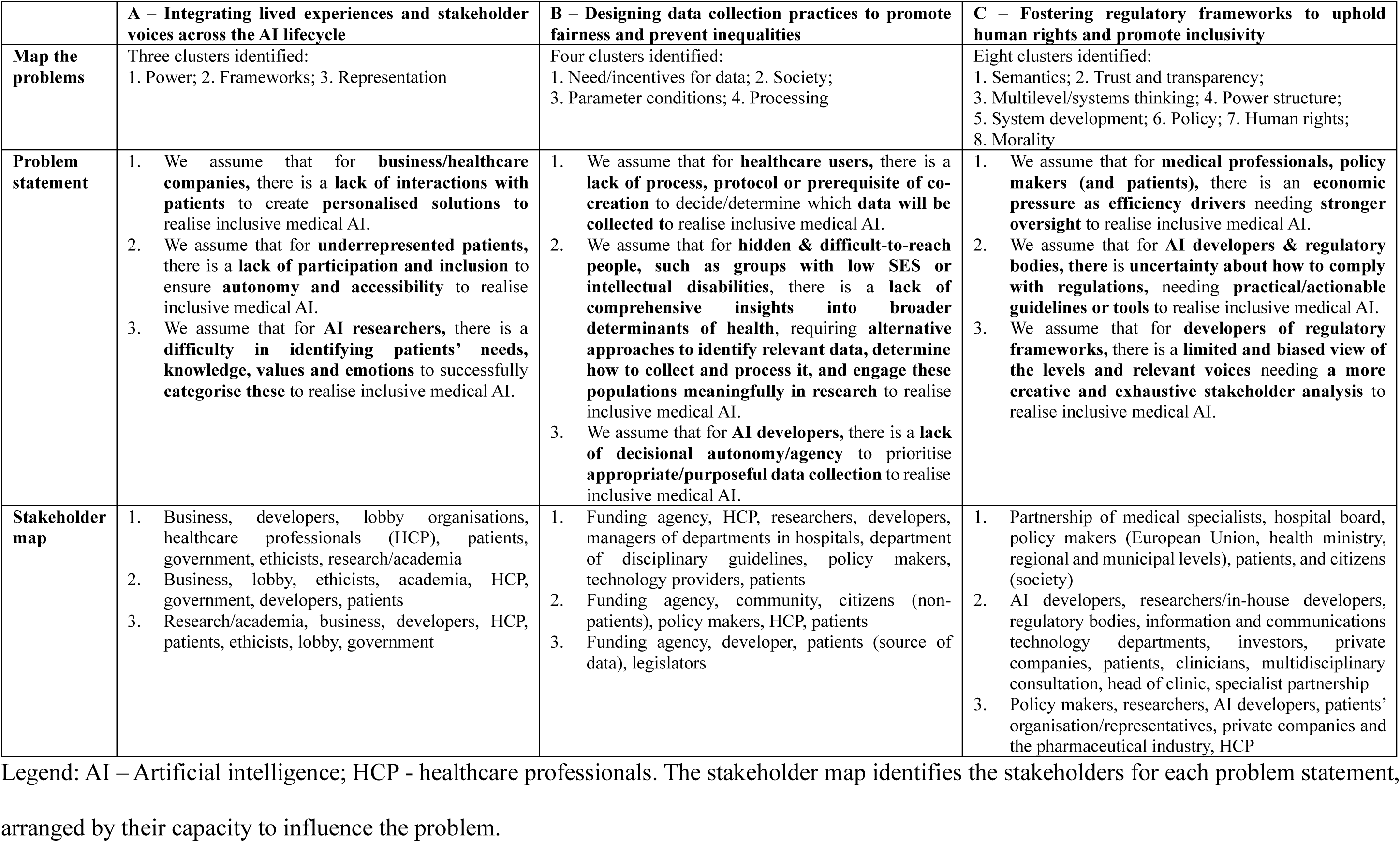
Problem statement for each challenge.

A central theme was the presence of structural and procedural barriers that disempower specific stakeholders (especially patients, disadvantaged and underserved groups, and individuals experiencing (multiple) layers of vulnerability, but also AI developers), leading to inequities in participation and decision-making. The three groups also identified the absence of integrated, practical, and inclusive frameworks as a significant limitation across all challenges: from a lack of methods to categorise patients’ values and needs (challenge A), to fragmented approaches to data co-creation and processing (challenge B), to uncertainty around regulatory compliance (challenge C). They also highlighted the need to improve the representation, engagement, and inclusion of diverse populations in AI design, development, validation, implementation, monitoring, and governance. Particular attention is required for underserved patients and people experiencing (multiple) layers of vulnerabilities to ensure that all voices are meaningfully considered.

Another common finding was the need for AI practices to be more responsive to lived experiences, contextual differences, and social determinants of health. During the stakeholder mapping exercise, participants identified the stakeholders most relevant to addressing each challenge, including policymakers, healthcare professionals (HCPs), developers, patients, industry, and researchers. While the exact configuration varied across challenges, there was a common emphasis on involving diverse stakeholders from different sectors and fostering partnerships (Table 2).

### Ideation

The *Ideation* stage led to the development of more than 180 solutions or requirements for inclusive medical AI. Through a thematic analysis and clustering conducted after the workshop, we synthesised these into eight key thematic clusters, which informed the development of the **PREFER-IT** framework (Fig 3).

- *P* stands for *Participatory and co-design approaches*. Participants consistently emphasised the importance of involving underserved and diverse communities, including patient representatives, throughout the entire AI lifecycle.
- *R* represents the need for *Representative and diverse data*. Participants stressed the importance of integrating data that accurately reflects the heterogeneity of patient populations and incorporates the lived experiences of patients.
- *E* refers to *Education and digital literacy* for enabling critical engagement with AI technologies. Participants highlighted the need for targeted educational initiatives to enhance the understanding of AI among patients, healthcare professionals, and the broader public, addressing both the potential benefits and risks.
- *F*, for *Fairness*, encapsulates the demand for the proactive integration of diversity, inclusivity, and equity considerations from the inception of AI development, and throughout all stages of the AI lifecycle.
- The second *E* denotes *Ethical and legal accountability*. Participants emphasised the need for clear governance structures and accountability mechanisms to address ethical, legal and regulatory challenges throughout the AI lifecycle.
- *R* also stands for *Real-world validation and feedback*. There was a broad consensus on the need for iterative refinement, testing and monitoring of AI tools across various settings, and enabling continuous user input for improvement.
- *I* represents *Inclusive communication*, focused on improving innovations that foster effective communication and decrease language and social barriers with and across diverse users and stakeholders.
- Finally, *T* signifies *Technical interoperability*. Participants highlighted the necessity of scalability and integration of AI models across heterogeneous infrastructures (e.g. data standards, electronic health record systems, and information system architectures), enabling compatibility across institutions and regions.

**Fig 3.**
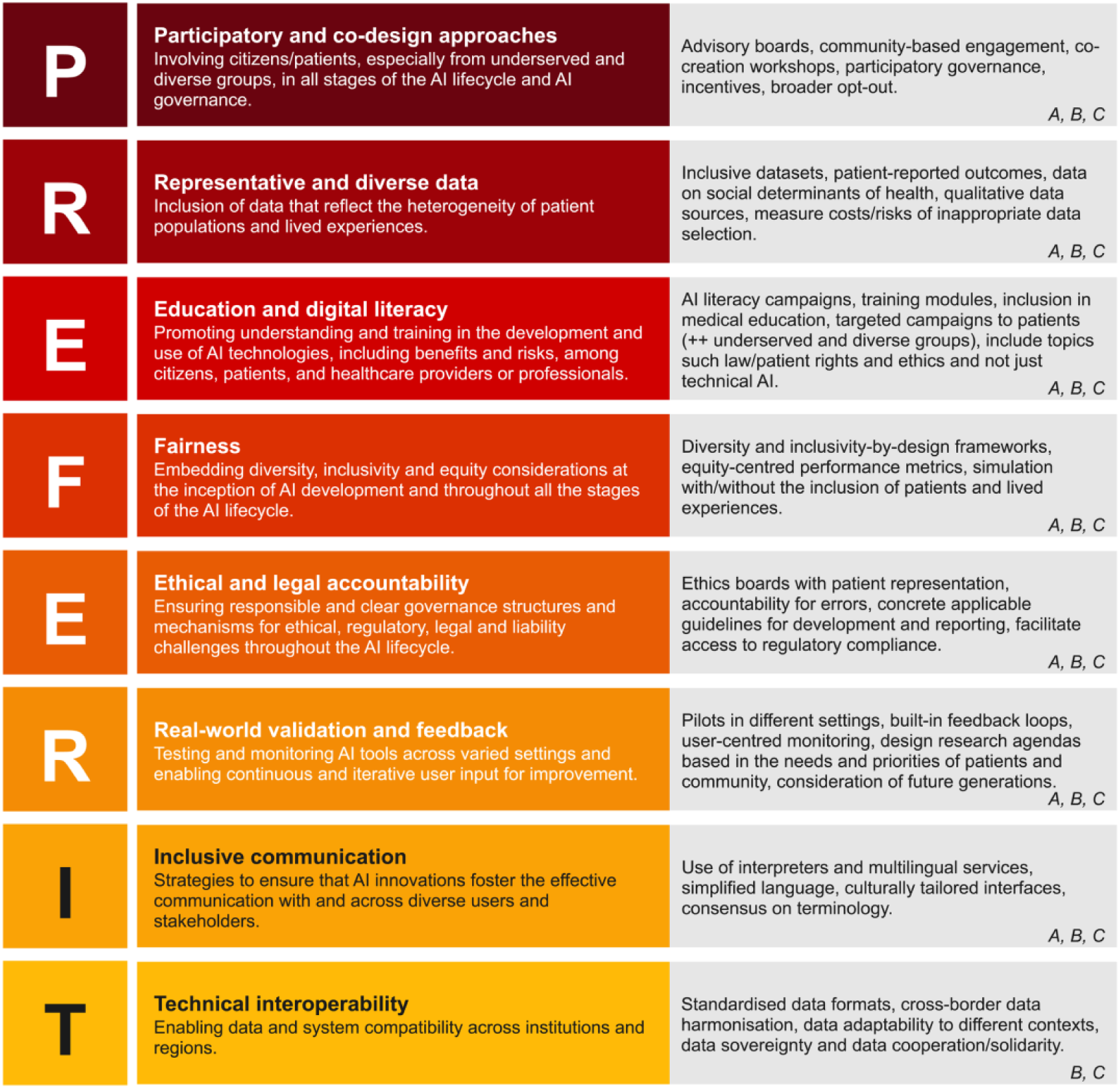
PREFER-IT framework. How PREFER-IT originated from the thematic clusters that emerged after ideation for solutions and requirements for inclusive medical AI, and clustering across the three challenges. In grey, the figure presents concrete solutions for implementing the PREFER-IT requirements across the workshop challenges (A, B, C).

### Prototype

Each group developed, refined and strengthened a set of three to four proposed solutions and requirements to advance inclusive medical AI (*Prototype* stage). The proposed solutions addressed various stages of the AI lifecycle and targeted both structural and procedural challenges (Table 3).

**Table 3.** Summary of proposed solutions by challenge.

|  | **A – Integrating lived experiences and stakeholder voices across the AI lifecycle** | **B – Designing data collection practices to promote fairness and prevent inequalities** | **C – Fostering regulatory frameworks to uphold human rights and promote inclusivity** |
| --- | --- | --- | --- |
| **Focus area** | Operationalisation of ethical and regulatory frameworks; participatory development | Equity in data governance; education and incentives | Regulatory support; community empowerment |
| **Proposed solutions** | 1. Develop a toolbox to support navigation of ethical and regulatory guidelines<br>2. Systematically involve diverse patient populations across AI lifecycle stages<br>3. Establish a European-level curriculum/training program for patient representatives in AI | 4. Improve transparency in data collection involving vulnerable populations<br>5. Embed data literacy in healthcare education<br>6. Create structural incentives (e.g., funding eligibility linked to data literacy)<br>7. Promote public and academic sector leadership in AI development | 8. Establish a multi-stakeholder advisory panel on inclusive AI<br>9. Create a compliance support tool for inclusivity alignment<br>10. Collect and disseminate grassroots success stories to foster mutual learning |
| **Intended impact** | - Support practical adoption of existing guidelines, particularly among developers, to create more impactful and responsible AI applications<br>- Promote sustained, meaningful stakeholder engagement<br>- Build capacity among patient representatives | - Foster fairness and equity in data practices<br>- Enhance critical data skills among professionals<br>- Reduce industry dependency<br>- Enable more inclusive AI innovation ecosystems | - Strengthen institutional guidance and accountability<br>- Support implementation of inclusivity principles<br>- Amplify local innovations and bottom-up engagement |

Group A focused on facilitating the practical implementation of existing ethical and regulatory guidelines by proposing the development of a comprehensive toolbox that breaks down how these frameworks, ranging from laws to inclusive AI principles in healthcare, can be applied throughout the AI development process. Additionally, the group highlighted the importance of systematically involving diverse patient populations throughout the AI development process and suggested establishing a European-level learning programme or course to train and empower patient representatives in AI. This programme would aim to build their literacy, capacity, and confidence to engage meaningfully and sustainably in AI design and governance processes. Another proposal involved organising bi-directional events, enabling patients and developers to gain a mutual understanding, fostering patient empowerment, and enhancing their agency in shaping AI solutions.

Group B proposed greater transparency and inclusiveness in data collection practices, particularly when involving vulnerable populations. They argued that data collection should become an explicit and mutual process between data collectors or domain experts (e.g., healthcare providers, researchers) and data subjects (e.g. patients, underserved groups), especially in encounters where such interactions are currently implicit or absent. The group advocated for embedding data literacy in healthcare education and establishing structural institutional incentives, such as requiring data literacy as a prerequisite for funding eligibility criteria within national funding agencies. Additionally, they highlighted the necessity of structural reform to reduce reliance on industry-led AI development by strengthening leadership in the public and academic sectors.

Group C emphasised the necessity of system change through far-reaching engagement with all the participatory stakeholders to foster inclusive AI development. They proposed the establishment of a multi-stakeholder advisory panel, composed of diverse stakeholder representatives, to be actively engaged throughout the development of regulatory frameworks, thereby ensuring that inclusivity principles are embedded across the AI lifecycle (focusing on the level of engagement). In parallel, they advocated for creating a compliance support tool integrating an interactive legal and ethical database with access to human legal expertise to assist developers and institutions in navigating regulatory and ethical requirements (focusing on the level of system change). They proposed systematically collecting and disseminating success stories from grassroots communities to support bottom-up engagement, which reflects diverse value priorities and local innovations (focusing on the levels of incentives). By increasing policymakers’ awareness of these community-driven practices, the initiative would seek to stimulate responsive policy changes, thereby fostering a dynamic interaction between bottom-up initiatives and top-down governance in AI innovation.

### Prioritise

At the *Prioritise* stage, participants identified and debated common themes and gaps for advancing inclusive medical AI. The findings reported here are based on the subsequent analysis of these discussions. (Table 4).

**Table 4.** Summary of shared priorities and gaps for advancing inclusive medical AI.

| Category | Description |
| --- | --- |
| **Shared Priorities** | - Development of support tools for guideline adoption and compliance<br>- Mechanisms for meaningful inclusion of diverse voices<br>- Integration of AI and data literacy into educational curricula<br>- Design of incentive structures to promote inclusivity<br>- Empowerment of marginalised communities<br>- Systemic, value-driven change<br>- Cross-boundary collaboration among disciplines, sectors, and stakeholders |
| **Gaps** | - Complexity and extended timelines of implementing inclusive, systemic solutions<br>- Difficulty in operationalising ethical principles amid rapid technological change and innovation<br>- Challenge of reconciling standardisation with value pluralism in diverse healthcare contexts<br>- Fragmentation of disciplinary, lifecycle, and stakeholder efforts<br>- Lack of proactive design for inclusivity (retrospective approaches dominate)<br>- Limited and inconsistent stakeholder interaction and knowledge exchange<br>- Insufficient patient involvement in innovation processes<br>- Weak mechanisms for influencing policy change |

Several overlapping themes emerged across these group discussions and the proposed solutions, reflecting a shared understanding of the systemic changes required to achieve inclusive medical AI. These included the development of support tools to assist in guideline adoption and compliance, mechanisms for the meaningful inclusion of diverse voices, and the integration of AI and data literacy into educational curricula. Other recurring themes involved designing incentive structures to encourage inclusive practices, such as linking funding to specific actions that promote data literacy and transparency. These incentives were seen as essential for empowering marginalised communities and driving broader systemic, value-driven change. The participants also recognised the fragmented nature of current efforts, particularly siloed approaches across disciplines, the uneven attention to different stages of the AI lifecycle, and the lack of integrated stakeholder engagement mechanisms. This underscored the importance of cross-boundary collaboration among disciplines, sectors, and stakeholders. Despite the promising nature of these proposals, participants acknowledged several persistent gaps that must be addressed to advance inclusive medical AI. One such gap is the inherent complexity and extended timelines involved in implementing inclusive solutions, particularly those that require systemic change, such as embedding participatory mechanisms across the AI lifecycle, reforming data governance structures, or aligning regulatory frameworks with human rights principles. These efforts often demand sustained stakeholder engagement, cross-sector coordination, and iterative testing, which can be resource-intensive and slow-moving.

Another gap raised by participants was safeguarding ethical principles during rapid technological change. While many stakeholders invoke ethical principles, such as fairness, transparency, or accountability, participants noted that these principles are often inconsistently applied or lack operational clarity in practice. The challenge lies not in the absence of ethical discourse, but in translating ethical principles into actionable, context-sensitive mechanisms that can keep pace with technological innovation. Participants emphasised the need for tools, training, and governance structures that support ethical alignment throughout the AI lifecycle, especially in high-stakes domains like healthcare.

A third gap involved reconciling standardisation with value pluralism. Participants noted that many AI systems in healthcare are designed for scalability and interoperability, which often necessitates standardised protocols and data formats. However, healthcare contexts are inherently diverse: patients, communities, and institutions may hold differing values, priorities, and definitions of equitable care. The challenge is not that unified systems are incompatible with diverse values, but that without deliberate design, standardisation can obscure or override context-specific needs. Participants emphasised the importance of embedding mechanisms for local adaptation, stakeholder input, and value-sensitive design to ensure that AI systems remain responsive to pluralistic healthcare environments.

Furthermore, participants emphasised the need for systems that are proactively designed to be inclusive from the beginning rather than retrospectively. They stressed the importance of sustained knowledge exchange and interaction among stakeholders, greater patient involvement in shaping innovation trajectories, and more effective mechanisms for influencing policy.

### PREFER-IT framework consolidation

To strengthen the practical value of PREFER-IT, we mapped it onto a dual-axis model (Fig 4), combining Zowghi and da Rimini’s structural layers of AI (humans, data, process, system, governance) [30] with the phases of the AI lifecycle (design, development, validation, implementation, monitoring) [2]. The *humans* pillar emphasises the centrality of stakeholder involvement and diverse representation. *Data* focuses on equitable and unbiased practices in the collection, labelling, modelling, and use. *Process* covers the activities across the lifecycle that lead to the delivery of an AI system. The *system* refers to the AI system itself, which must be evaluated, tested, and monitored in context. Finally, the *governance* pillar comprises the structures, processes, and regulatory frameworks that ensure compliance and ethical alignment.

**Fig 4.**
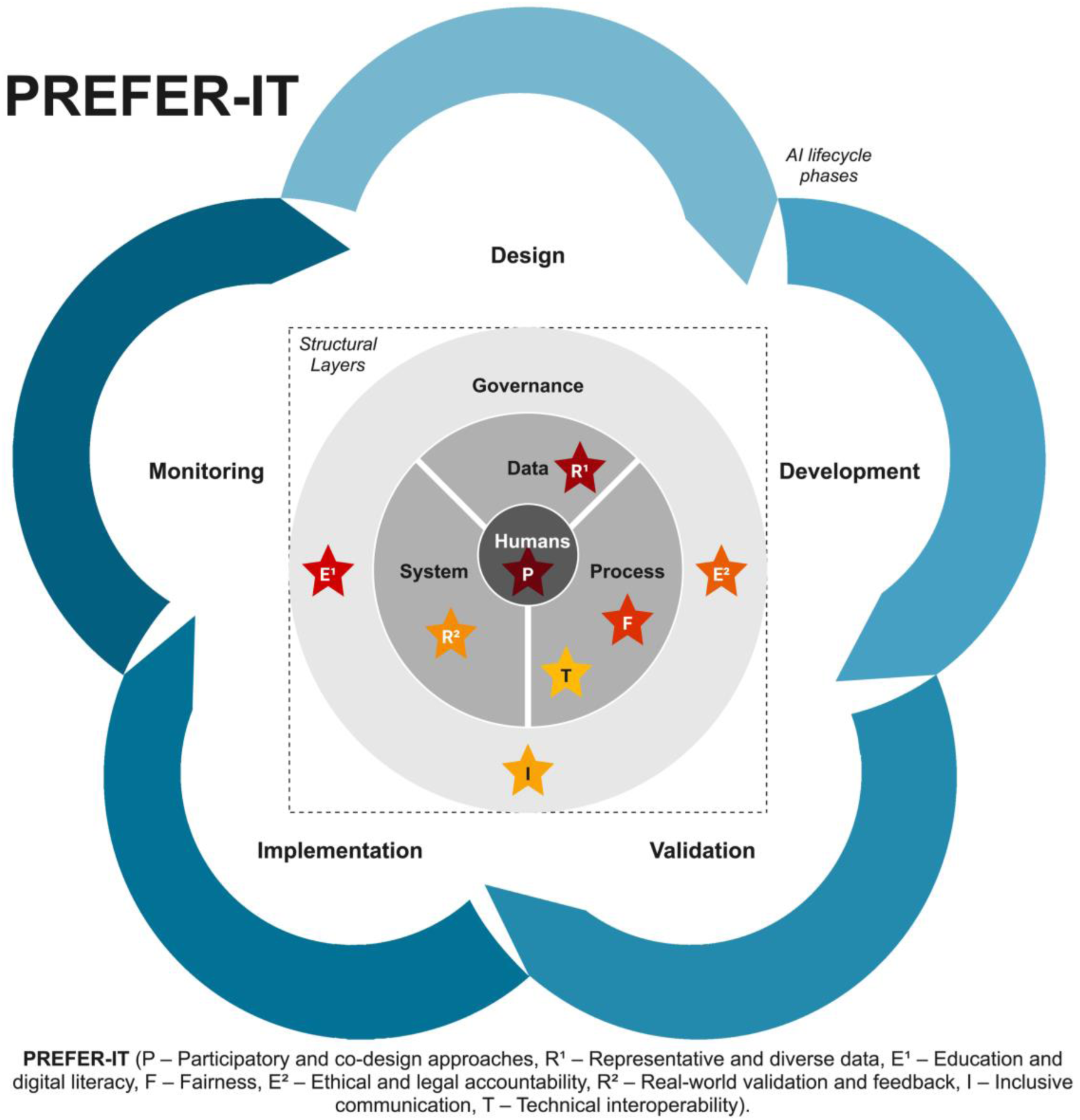
Implementation of the PREFER-IT framework across structural layers and the AI lifecycle.

This approach situates PREFER-IT elements as requirements that can orient and sustain inclusivity across different dimensions of medical AI.

Importantly, the boundaries of PREFER-IT within this dual axis are not meant to be static, but are intentionally flexible. This flexibility reflects the reality that many requirements intersect across different AI structural layers and lifecycle phases, underscoring the interdependent and systemic nature of inclusivity in AI.

For simplification purposes, in Fig 4 each component of the PREFER-IT framework aligns with specific structural layers and phases of the medical AI lifecycle. For instance, Participatory and co-design approaches (P) are essential throughout all stages of the AI lifecycle and must actively involve all stakeholders (what is labelled as “humans”). Representative and diverse data (R) plays a crucial role within the data layer, particularly during the design phase. Elements such as Education and digital literacy (E), Ethical and legal accountability (E), and Inclusive communication (I) are fundamental governance considerations that span the entire AI lifecycle. Meanwhile, Fairness (F) and Technical interoperability (T) are core to the process layer and should be prioritised during development and validation. Finally, Real-world validation and feedback (R) are vital in any AI system, ensuring its adaptation to evolving socio-technical contexts, and should be considered during the validation, implementation and monitoring stages.

To demonstrate a proof of concept, we mapped the representative prototype solutions developed during the in-person workshop (from Table 3) onto the dual-axis of PREFER-IT (Fig 5). This exercise highlights how the framework can be applied in practice with concrete solutions across the AI ecosystem.

**Fig 5.**
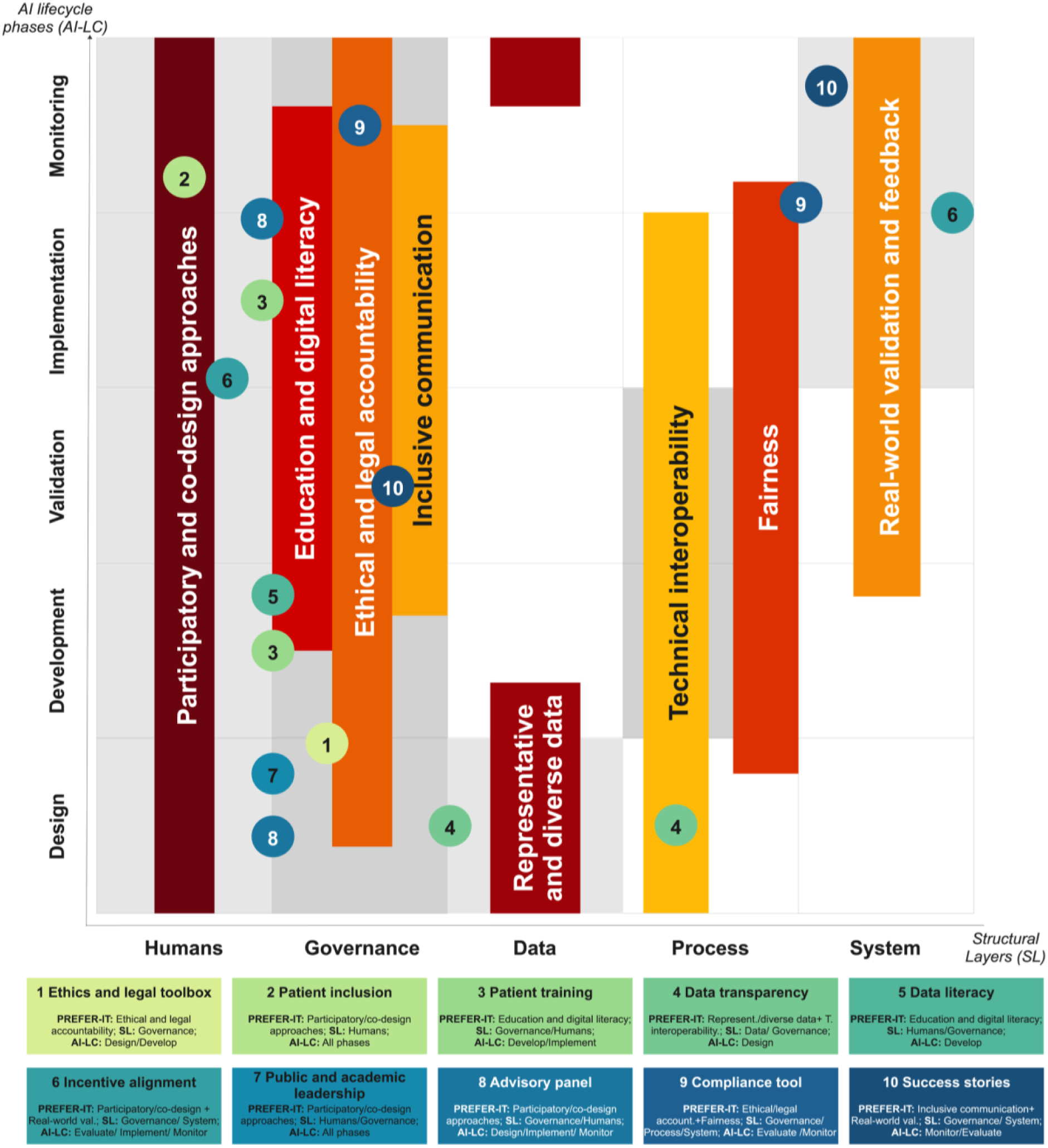
Operationalising PREFER-IT. Mapping of the prototype solutions co-created during the workshop (from Table 3) across the dual-axis of PREFER-IT (AI structural layers and lifecycle stages).

A single solution can justifiably be placed in different locations depending on the perspective of the team or stakeholder group conducting the mapping. Rather than undermining the map’s purpose, this adaptability reinforces its value as a practical tool for navigating complexity. The value of this exercise is not to produce a definitive categorisation, but to use PREFER-IT as a guiding framework that ensures inclusivity remains a core consideration throughout the design, development, validation, implementation, monitoring, and governance of AI in healthcare.

## Discussion

This paper presents PREFER-IT, a set of co-created foundational requirements developed through a transdisciplinary workshop to provide an operational framework for realising inclusivity throughout the AI lifecycle and governance in healthcare. By engaging a diverse group of stakeholders in a structured co-creation process, we translated lived experiences and expert insights into actionable requirements for inclusive medical AI.

The workshop combined a preparatory PPI meeting and survey, a consensus-building exercise, and design thinking methods, including patient personas and journey mapping with the active involvement of patient experts. This structured process progressively moved from the broad challenge of inclusive medical AI to a set of concrete, experience-informed solutions and requirements, eventually culminating in the development of the PREFER-IT framework.

While existing frameworks often address fairness, trustworthiness or ethics in medical AI [2,9,18,25,43–46], they generally remain conceptual or focus on technical dimensions. A unified, actionable framework for AI inclusivity, particularly within healthcare, remains absent, at least to our knowledge. PREFER-IT is a first step to respond to this gap by offering both a conceptual and applied foundation to guide the inclusive design, development, validation, implementation, monitoring, and governance of medical AI.

The co-creation process involved a small number of participants, providing an in-depth and explorative knowledge and exchange that generated valuable insights. The diversity of expertise and richness of perspectives provide a robust qualitative foundation for further empirical investigation. The co-created solutions, grounded in patient voices and lived experiences, particularly emphasised the human and governance dimensions of AI, reflecting the workshop’s interdisciplinary composition across social sciences, ethics, and health policy. However, the framework remains adaptable to technical domains and can guide integration at the data, process, and system levels of medical AI. While the discussions were primarily situated within a European context, this framing offers a strong basis for extending and validating the framework in other settings. A possible next step could be to test and adapt the framework across diverse contexts to capture a broader range of epistemic perspectives and societal concerns, ensuring its global relevance and applicability.

The workshop also revealed the importance of authentic participation. While patient involvement enriched the framework, it also highlighted risks of tokenism. Similar tensions have been observed in participatory AI design more broadly, where researchers and practitioners often struggle to reconcile participatory ambitions with practical constraints such as time, resources, and institutional norms [47]. To navigate these challenges, teams frequently rely on proxies (like stand-ins for stakeholders) that may inadvertently reinforce existing power asymmetries rather than dismantle them [47]. This underscores the need for co-creation to extend beyond isolated workshops toward sustained, reflexive engagement mechanisms that distribute decision-making authority and epistemic legitimacy. Without such safeguards, inclusion risks being symbolic rather than transformative.

Participants also identified several persistent gaps for realising inclusive medical AI. Rather than obstacles, these can be reframed as priority areas where PREFER-IT should be actively applied or advocated. For instance, the call for support tools to facilitate guideline adoption and compliance highlights the need for stronger public engagement in shaping and implementing AI regulation, as illustrated by the Algorithm Register of the Dutch government [48]. At the policy level, incorporating PREFER-IT principles into funding criteria, regulatory assessments, and institutional governance could help move inclusion from aspiration to routine practice. This approach would shape AI medical inclusivity in *what* is developed, *how* it is developed, *who* develops it, and *how* it is evaluated and refined over time.

Likewise, the demand for educational and capacity-building initiatives, such as patient training programs or education tools, aligns with ongoing policy efforts, such as those outlined in the AI Act [49], to support sustained implementation of PREFER-IT. Capacity-building should also target AI developers and healthcare professionals to bridge disciplinary gaps. As AI researchers often lack medical expertise, collaboration with clinicians and patients is crucial for understanding the clinical context and translating patient needs into technical design requirements.

The more systemic concerns, including implementation complexity, safeguarding ethics, and balancing diverse values, highlight that PREFER-IT should not be approached as a simple checklist. Instead, it needs to be an adaptable infrastructure spanning the entire AI lifecycle and its various structural layers. The core challenge lies not in the absence of ethical discourse but in translating principles into actionable, context-sensitive mechanisms that can keep pace with technological innovation. Viewing inclusivity as a behavioural process that requires capability, opportunity, and motivation could strengthen the framework’s practical applicability and promote organisational cultures that sustain inclusive AI development. [50]

Looking ahead, the next steps involve translating PREFER-IT from a co-created framework into a living infrastructure for inclusive medical AI. This will require ongoing engagement with diverse stakeholders, including patients with intersectional vulnerabilities, regulators, and insurers. It also involves iterative testing in real-world projects, systematic evaluation across healthcare contexts, and continuous refinement based on user and patient feedback. Through this operationalisation, PREFER-IT aims to advance AI in healthcare that is not only effective but also equitable, ethical, legal, and socially responsible [51].

The co-creation process highlighted that realising inclusivity in medical AI requires more than just design features, depending on systemic enablers across multiple levels. In line with Wang and Blok’s multi-level framework [25], PREFER-IT can be operationalised at the micro-level (individual AI issues), meso-level (organisational and systemic issues), and macro-level (philosophical issues). For example, policymakers can mandate transparency and equity audits; developers can integrate inclusion-by-design and participatory testing; researchers and clinicians can promote representative study designs; and patients and communities can co-define research priorities and evaluation metrics. The PREFER-IT framework could provide a basis for coordinating these actions across roles and governance levels.

Finally, our research suggests a broader reframing: from an *innovation-first* logic to an *inclusive-by-design* paradigm. This shift positions inclusivity as a constitutive element of medical AI, embedded not only in outcomes but also in processes, governance structures, and power relations. By doing so, PREFER-IT helps address epistemic hierarchies in knowledge generation, challenging the dominance of quantitative and technical evidence and elevating the value of qualitative insights, experiential knowledge, and patient voices in shaping AI development.

Through the development of PREFER-IT, we offer both a conceptual contribution and a practical resource to support stakeholders in realising inclusivity throughout the AI lifecycle in healthcare. The framework builds on, but also extends beyond, existing frameworks by positioning inclusion not as a peripheral ethical concern but as a core design principle, rooted in equity, stakeholder engagement, and governance accountability.

Crucially, this process revealed that advancing inclusion in medical AI is not merely a matter of technological or design choices. It requires reconfiguring institutional logics, incentive structures, and policy environments. The PREFER-IT framework captures these multi-level dynamics and provides a framework for coordinated action. By explicitly incorporating the roles of patients and communities, it helps bridge the gap between AI development and implementation and the lived realities of those most affected by it.

Ultimately, this process challenges dominant epistemic practices and paradigms in medical innovation, which often prioritise efficiency and technical advancement over equity and participation. Co-creation, as adopted here, is not merely a methodological technique but a stance that recognises the value of inclusion. It shifts the focus from innovation *for* to innovation *with* communities.

## Author Contributions

Conceptualisation: PPF, SSL, WB, SdK, CL, LB, MP; Data Curation: PPF, SSL, WB, LB; Investigation: PPF, SSL, WB, JG, LB, MP; Methodology: PPF, SSL, WB, JG, LB, MP; Supervision: SdK, CL, MP; Visualisation: PPF; Writing – Original Draft Preparation: PPF; Writing – Review & Editing: all the authors

## Acknowledgments

The authors would like to thank the Lorentz Centre (Leiden, The Netherlands) for hosting the workshop “Inclusive medical AI: Exploring Diverse Perspectives” from February 10 to 14, 2025.

## Supporting information captions

**S1 Fig.**
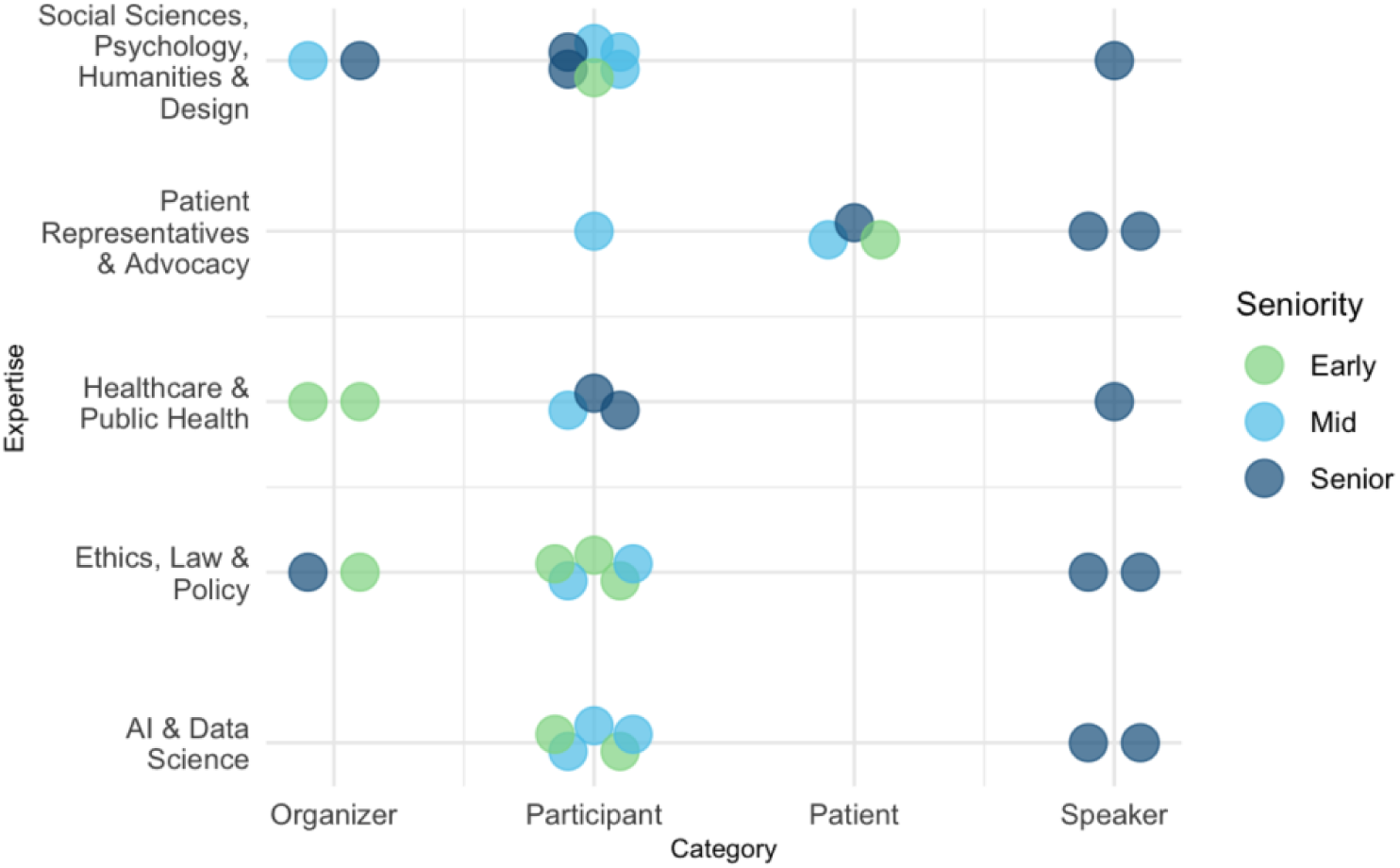
Field of expertise and seniority of workshop contributors by category. Distribution of experts involved in the workshop on "Inclusive Medical AI: Exploring Diverse Perspectives" according to category, field of expertise, and seniority.

**S2 Fig.**
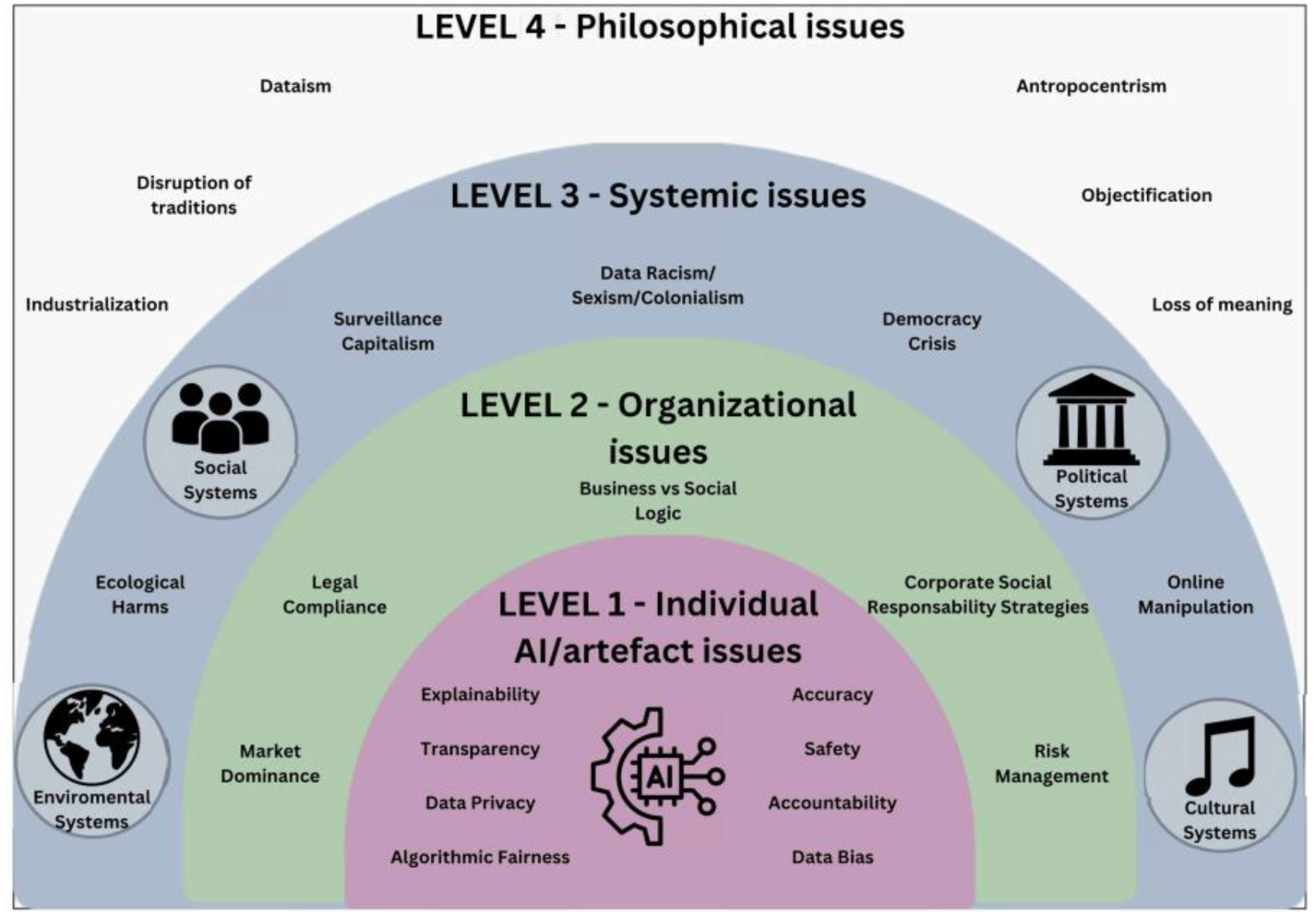
Relevant socio-ethical issues of inclusive medical AI, adapted from Wang and Blok’s multi-level framework. The framework was used to map the identified challenges of inclusive medical AI and their corresponding requirements across different levels.

**S1 Table.** Results from Pol.is.

| ID statement | Statement | Overall (N=27) |  |  |  |  | Group A (N=10) |  |  |  |  | Group B (N=4) |  |  |  |  | Group C (N=5) |  |  |  |  | Group D (N=8) |  |  |  |  |
| --- | --- | --- | --- | --- | --- | --- | --- | --- | --- | --- | --- | --- | --- | --- | --- | --- | --- | --- | --- | --- | --- | --- | --- | --- | --- | --- |
|  |  | Total votes | Agree | Disagree | Pass | No vote | Total votes | Agree | Disagree | Pass | No vote | Total votes | Agree | Disagree | Pass | No vote | Total votes | Agree | Disagree | Pass | No vote | Total votes | Agree | Disagree | Pass | No vote |
|  |  | N | N (%) | N (%) | N (%) | N | N | N (%) | N (%) | N (%) | N | N | N (%) | N (%) | N (%) | N | N | N (%) | N (%) | N (%) | N | N | N (%) | N (%) | N (%) | N |
| 0 | Developing methods to minimize biases in AI systems. | 27 | 18 | 4 | 5 | 0 | 10 | 5 | 2 | 3 | 0 | 4 | 4 | 0 | 0 | 0 | 5 | 5 | 0 | 0 | 0 | 8 | 4 | 2 | 2 | 0 |
|  |  |  | 66,7% | 14,8% | 18,5% |  |  | 50,0% | 20,0% | 30,0% |  |  | 100,0% | 0,0% | 0,0% |  |  | 100,0% | 0,0% | 0,0% |  |  | 50,0% | 25,0% | 25,0% |  |
| 1 | Understanding the effects of bias on AI outputs, such as unfair recommendations. | 27 | 15 | 3 | 9 | 0 | 10 | 2 | 3 | 5 | 0 | 4 | 4 | 0 | 0 | 0 | 5 | 5 | 0 | 0 | 0 | 8 | 4 | 0 | 4 | 0 |
|  |  |  | 55,6% | 11,1% | 33,3% |  |  | 20,0% | 30,0% | 50,0% |  |  | 100,0% | 0,0% | 0,0% |  |  | 100,0% | 0,0% | 0,0% |  |  | 50,0% | 0,0% | 50,0% |  |
| 2 | Including diverse stakeholder voices throughout the AI lifecycle. | 26 | 21 | 1 | 4 | 1 | 9 | 7 | 0 | 2 | 1 | 4 | 3 | 1 | 0 | 0 | 5 | 5 | 0 | 0 | 0 | 8 | 6 | 0 | 2 | 0 |
|  |  |  | 80,8% | 3,8% | 15,4% |  |  | 77,8% | 0,0% | 22,2% |  |  | 75,0% | 25,0% | 0,0% |  |  | 100,0% | 0,0% | 0,0% |  |  | 75,0% | 0,0% | 25,0% |  |
| 3 | Including the lived experiences of diverse groups in AI design and implementation. | 27 | 22 | 0 | 5 | 0 | 10 | 8 | 0 | 2 | 0 | 4 | 4 | 0 | 0 | 0 | 5 | 5 | 0 | 0 | 0 | 8 | 5 | 0 | 3 | 0 |
|  |  |  | 81,5% | 0,0% | 18,5% |  |  | 80,0% | 0,0% | 20,0% |  |  | 100,0% | 0,0% | 0,0% |  |  | 100,0% | 0,0% | 0,0% |  |  | 62,5% | 0,0% | 37,5% |  |
| 4 | Developing regulatory frameworks that address inclusivity in medical AI. | 26 | 15 | 4 | 7 | 1 | 9 | 4 | 3 | 2 | 1 | 4 | 1 | 1 | 2 | 0 | 5 | 5 | 0 | 0 | 0 | 8 | 5 | 0 | 3 | 0 |
|  |  |  | 57,7% | 15,4% | 26,9% |  |  | 44,4% | 33,3% | 22,2% |  |  | 25,0% | 25,0% | 50,0% |  |  | 100,0% | 0,0% | 0,0% |  |  | 62,5% | 0,0% | 37,5% |  |
| 5 | Creating regulatory frameworks interpretable for all stakeholders. | 27 | 10 | 9 | 8 | 0 | 10 | 0 | 6 | 4 | 0 | 4 | 0 | 2 | 2 | 0 | 5 | 5 | 0 | 0 | 0 | 8 | 5 | 1 | 2 | 0 |
|  |  |  | 37,0% | 33,3% | 29,6% |  |  | 0,0% | 60,0% | 40,0% |  |  | 0,0% | 50,0% | 50,0% |  |  | 100,0% | 0,0% | 0,0% |  |  | 62,5% | 12,5% | 25,0% |  |
| 6 | Ensuring training datasets represent diverse groups. | 27 | 18 | 3 | 6 | 0 | 10 | 5 | 2 | 3 | 0 | 4 | 2 | 1 | 1 | 0 | 5 | 5 | 0 | 0 | 0 | 8 | 6 | 0 | 2 | 0 |
|  |  |  | 66,7% | 11,1% | 22,2% |  |  | 50,0% | 20,0% | 30,0% |  |  | 50,0% | 25,0% | 25,0% |  |  | 100,0% | 0,0% | 0,0% |  |  | 75,0% | 0,0% | 25,0% |  |
| 7 | Assessing whether current data collection practices exacerbate inequalities. | 27 | 20 | 3 | 4 | 0 | 10 | 5 | 3 | 2 | 0 | 4 | 4 | 0 | 0 | 0 | 5 | 5 | 0 | 0 | 0 | 8 | 6 | 0 | 2 | 0 |
|  |  |  | 74,1% | 11,1% | 14,8% |  |  | 50,0% | 30,0% | 20,0% |  |  | 100,0% | 0,0% | 0,0% |  |  | 100,0% | 0,0% | 0,0% |  |  | 75,0% | 0,0% | 25,0% |  |
| 8 | Investigating the societal impact of AI systems on diverse groups. | 27 | 17 | 5 | 5 | 0 | 10 | 4 | 4 | 2 | 0 | 4 | 4 | 0 | 0 | 0 | 5 | 5 | 0 | 0 | 0 | 8 | 4 | 1 | 3 | 0 |
|  |  |  | 63,0% | 18,5% | 18,5% |  |  | 40,0% | 40,0% | 20,0% |  |  | 100,0% | 0,0% | 0,0% |  |  | 100,0% | 0,0% | 0,0% |  |  | 50,0% | 12,5% | 37,5% |  |
| 9 | Continuous evaluation of AI across diverse groups. | 26 | 13 | 4 | 9 | 1 | 9 | 4 | 3 | 2 | 1 | 4 | 1 | 1 | 2 | 0 | 5 | 5 | 0 | 0 | 0 | 8 | 3 | 0 | 5 | 0 |
|  |  |  | 50,0% | 15,4% | 34,6% |  |  | 44,4% | 33,3% | 22,2% |  |  | 25,0% | 25,0% | 50,0% |  |  | 100,0% | 0,0% | 0,0% |  |  | 37,5% | 0,0% | 62,5% |  |
| 10 | Identifying how medical AI affects human rights of diverse groups. | 27 | 10 | 9 | 8 | 0 | 10 | 0 | 7 | 3 | 0 | 4 | 1 | 1 | 2 | 0 | 5 | 5 | 0 | 0 | 0 | 8 | 4 | 1 | 3 | 0 |
|  |  |  | 37,0% | 33,3% | 29,6% |  |  | 0,0% | 70,0% | 30,0% |  |  | 25,0% | 25,0% | 50,0% |  |  | 100,0% | 0,0% | 0,0% |  |  | 50,0% | 12,5% | 37,5% |  |
| 11 | Identifying how medical AI affects public and personal values of diverse groups. | 27 | 15 | 3 | 9 | 0 | 10 | 4 | 3 | 3 | 0 | 4 | 4 | 0 | 0 | 0 | 5 | 4 | 0 | 1 | 0 | 8 | 3 | 0 | 5 | 0 |
|  |  |  | 55,6% | 11,1% | 33,3% |  |  | 40,0% | 30,0% | 30,0% |  |  | 100,0% | 0,0% | 0,0% |  |  | 80,0% | 0,0% | 20,0% |  |  | 37,5% | 0,0% | 62,5% |  |
| 12 | Incorporating AI advancements in healthcare context. | 26 | 10 | 9 | 7 | 1 | 9 | 0 | 6 | 3 | 1 | 4 | 4 | 0 | 0 | 0 | 5 | 4 | 0 | 1 | 0 | 8 | 2 | 3 | 3 | 0 |
|  |  |  | 38,5% | 34,6% | 26,9% |  |  | 0,0% | 66,7% | 33,3% |  |  | 100,0% | 0,0% | 0,0% |  |  | 80,0% | 0,0% | 20,0% |  |  | 25,0% | 37,5% | 37,5% |  |
| 13 | Monitoring and detecting problems and risks in AI systems before their release. | 27 | 17 | 4 | 6 | 0 | 10 | 4 | 3 | 3 | 0 | 4 | 3 | 0 | 1 | 0 | 5 | 4 | 1 | 0 | 0 | 8 | 6 | 0 | 2 | 0 |
|  |  |  | 63,0% | 14,8% | 22,2% |  |  | 40,0% | 30,0% | 30,0% |  |  | 75,0% | 0,0% | 25,0% |  |  | 80,0% | 20,0% | 0,0% |  |  | 75,0% | 0,0% | 25,0% |  |
| 14 | Collaborating between different scientific disciplines. | 27 | 14 | 4 | 9 | 0 | 10 | 2 | 3 | 5 | 0 | 4 | 4 | 0 | 0 | 0 | 5 | 5 | 0 | 0 | 0 | 8 | 3 | 1 | 4 | 0 |
|  |  |  | 51,9% | 14,8% | 33,3% |  |  | 20,0% | 30,0% | 50,0% |  |  | 100,0% | 0,0% | 0,0% |  |  | 100,0% | 0,0% | 0,0% |  |  | 37,5% | 12,5% | 50,0% |  |
| 15 | Using a systems approach. | 25 | 7 | 4 | 14 | 2 | 8 | 2 | 3 | 3 | 2 | 4 | 4 | 0 | 0 | 0 | 5 | 0 | 1 | 4 | 0 | 8 | 1 | 0 | 7 | 0 |
|  |  |  | 28,0% | 16,0% | 56,0% |  |  | 25,0% | 37,5% | 37,5% |  |  | 100,0% | 0,0% | 0,0% |  |  | 0,0% | 20,0% | 80,0% |  |  | 12,5% | 0,0% | 87,5% |  |
| 16 | As a matter of principle, I don't think an AI tool should be used in the process of determining which topics are most pertinent. | 25 | 4 | 16 | 5 | 2 | 9 | 1 | 6 | 2 | 1 | 4 | 0 | 3 | 1 | 0 | 4 | 1 | 3 | 0 | 1 | 8 | 2 | 4 | 2 | 0 |
|  |  |  | 16,0% | 64,0% | 20,0% |  |  | 11,1% | 66,7% | 22,2% |  |  | 0,0% | 75,0% | 25,0% |  |  | 25,0% | 75,0% | 0,0% |  |  | 25,0% | 50,0% | 25,0% |  |
| 17 | Inventory and development of methods to include people in process of AI development and implementation | 22 | 12 | 3 | 7 | 5 | 8 | 4 | 1 | 3 | 2 | 4 | 1 | 2 | 1 | 0 | 2 | 1 | 0 | 1 | 3 | 8 | 6 | 0 | 2 | 0 |
|  |  |  | 54,5% | 13,6% | 31,8% |  |  | 50,0% | 12,5% | 37,5% |  |  | 25,0% | 50,0% | 25,0% |  |  | 50,0% | 0,0% | 50,0% |  |  | 75,0% | 0,0% | 25,0% |  |
| 18 | Adress AI inclusivity in different stages of the AI lifecycle | 22 | 16 | 1 | 5 | 5 | 8 | 6 | 1 | 1 | 2 | 4 | 3 | 0 | 1 | 0 | 2 | 2 | 0 | 0 | 3 | 8 | 5 | 0 | 3 | 0 |
|  |  |  | 72,7% | 4,5% | 22,7% |  |  | 75,0% | 12,5% | 12,5% |  |  | 75,0% | 0,0% | 25,0% |  |  | 100,0% | 0,0% | 0,0% |  |  | 62,5% | 0,0% | 37,5% |  |
| 19 | Adress AI inclusivity at different 'system' levels (micro, meso, macro) | 22 | 16 | 1 | 5 | 5 | 8 | 6 | 1 | 1 | 2 | 4 | 3 | 0 | 1 | 0 | 2 | 1 | 0 | 1 | 3 | 8 | 6 | 0 | 2 | 0 |
|  |  |  | 72,7% | 4,5% | 22,7% |  |  | 75,0% | 12,5% | 12,5% |  |  | 75,0% | 0,0% | 25,0% |  |  | 50,0% | 0,0% | 50,0% |  |  | 75,0% | 0,0% | 25,0% |  |
| 20 | Develop methods/frameworks to promote inclusivity for medical experts working with AI to increase safe adoption (e.g. AI literacy) | 20 | 12 | 4 | 4 | 7 | 8 | 5 | 1 | 2 | 2 | 4 | 1 | 2 | 1 | 0 | 2 | 1 | 0 | 1 | 3 | 6 | 5 | 1 | 0 | 2 |
|  |  |  | 60,0% | 20,0% | 20,0% |  |  | 62,5% | 12,5% | 25,0% |  |  | 25,0% | 50,0% | 25,0% |  |  | 50,0% | 0,0% | 50,0% |  |  | 83,3% | 16,7% | 0,0% |  |
| 21 | Make sure AI is at the service of patients | 19 | 9 | 3 | 7 | 8 | 7 | 3 | 2 | 2 | 3 | 4 | 3 | 0 | 1 | 0 | 2 | 2 | 0 | 0 | 3 | 6 | 1 | 1 | 4 | 2 |
|  |  |  | 47,4% | 15,8% | 36,8% |  |  | 42,9% | 28,6% | 28,6% |  |  | 75,0% | 0,0% | 25,0% |  |  | 100,0% | 0,0% | 0,0% |  |  | 16,7% | 16,7% | 66,7% |  |
| 22 | When developing and deploying healthcare AI, patients' rights should be taken into account. | 6 | 4 | 1 | 1 | 21 | 1 | 0 | 1 | 0 | 9 | 0 | 0 | 0 | 0 | 4 | 1 | 1 | 0 | 0 | 4 | 4 | 3 | 0 | 1 | 4 |
|  |  |  | 66,7% | 16,7% | 16,7% |  |  | 0,0% | 100,0% | 0,0% |  |  | NA | NA | NA |  |  | 100,0% | 0,0% | 0,0% |  |  | 75,0% | 0,0% | 25,0% |  |

Legend: The statements highlighted in grey represent those with the highest levels of agreement (defined as having over 66% of participants selecting “Agree” across the entire sample). Participants were asked to vote on the statements, considering what they felt were the most significant challenges we should focus on during the five-day in-person workshop. This means that when participants choose “Disagree” or “Pass”, it is not necessarily because they disagree or are unsure about the statement, but because they felt it was not as big a priority to focus on the workshop as other statements. Among the 27 participants, four distinct opinion groups emerged in Pol.is. These groups were defined by two criteria: first, members within each group exhibited similar voting patterns across multiple statements; second, their voting behaviour was consistently different from that of participants in the other groups. Pol.is uses the k-means clustering algorithm to group participants into clusters and the silhouette coefficient to select an optimal number of clusters.

**S3 Fig.**
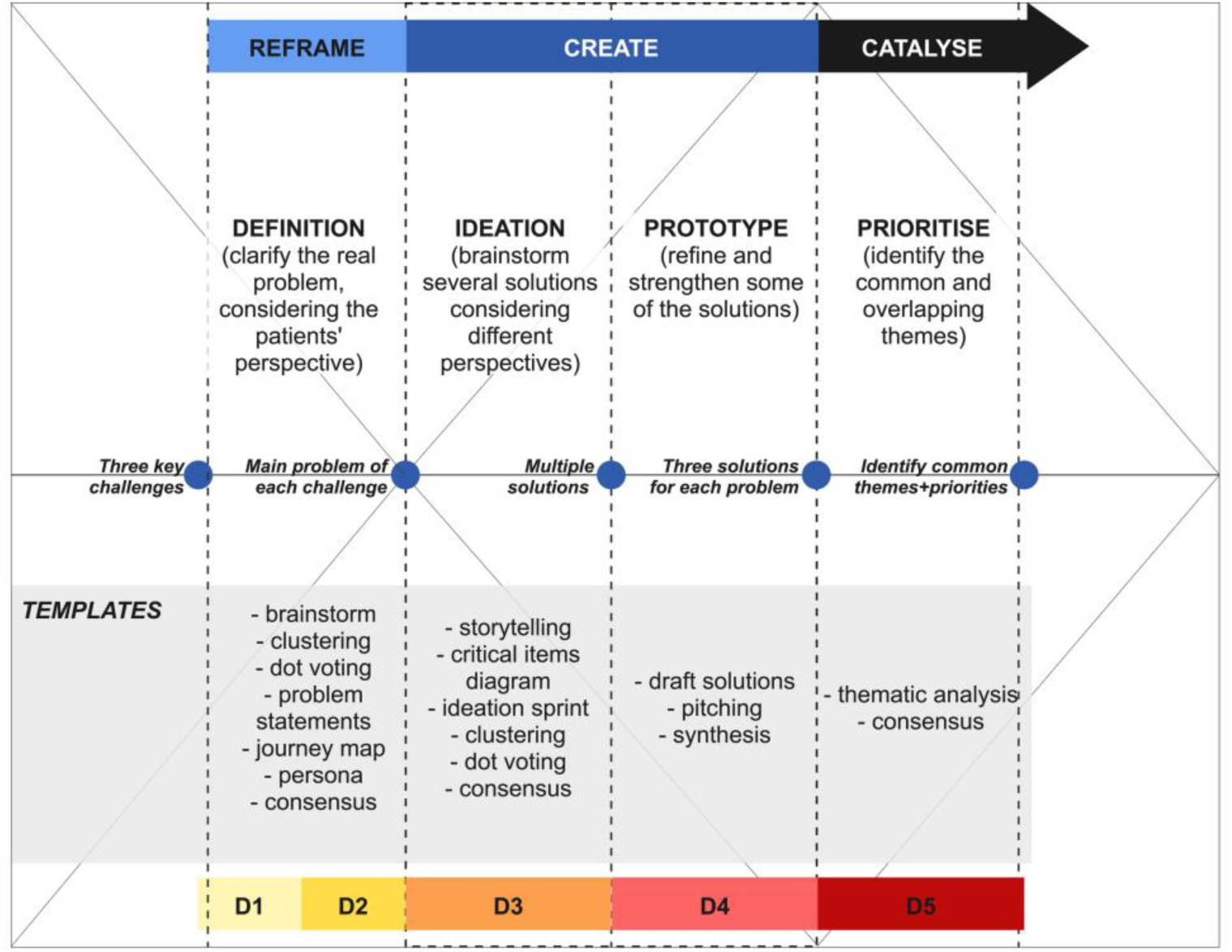
Prepared templates. Templates prepared for each day of the workshop, based on the double diamond model and the innovation and systemic design frameworks, [36–38]

**S4 Fig.**
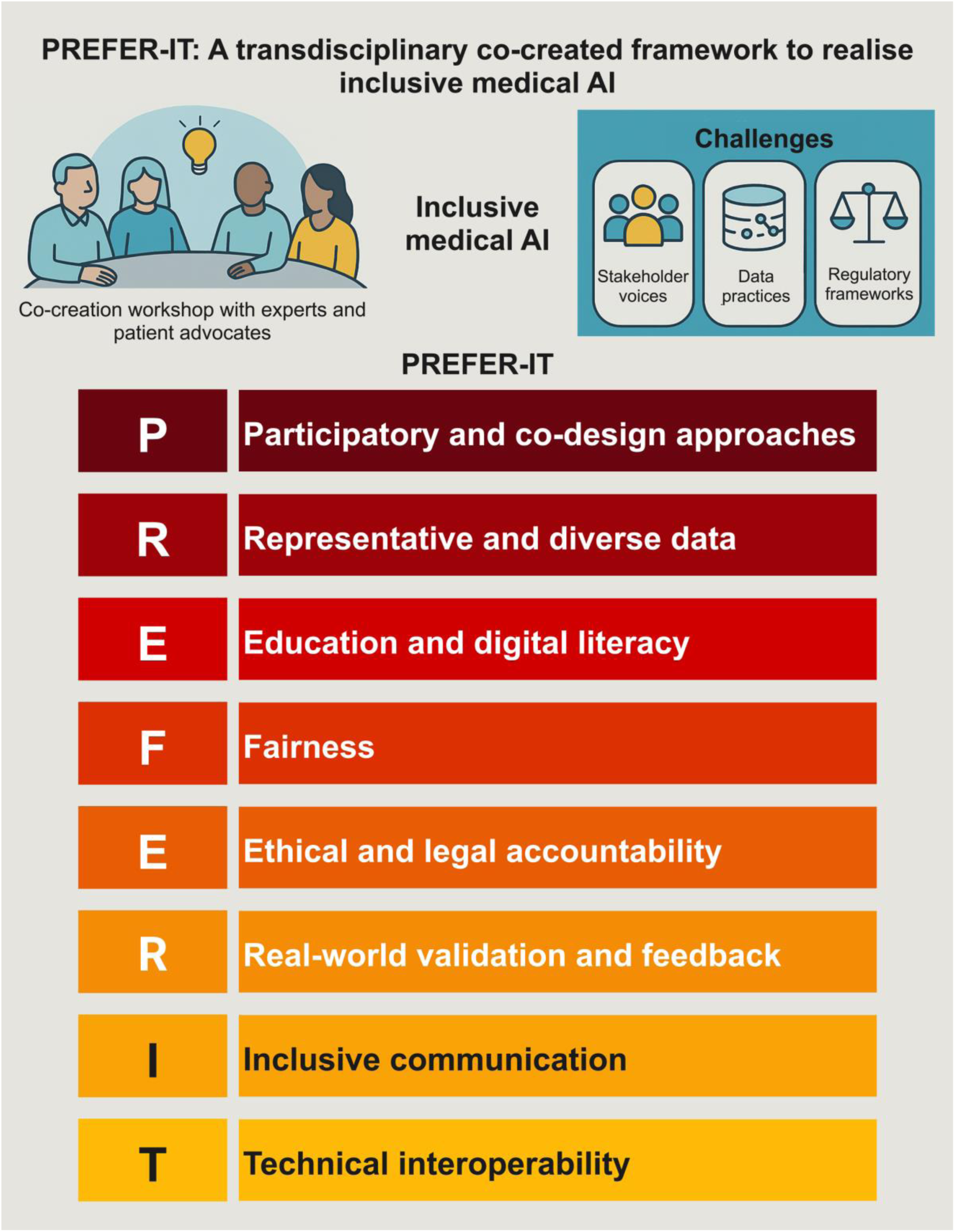
Graphic abstract.

## Data Availability

The data collected and analysed during the workshop “Inclusive Medical AI: Exploring Diverse Perspectives” consist of anonymised workshop notes, co-created personas, and anonymous patient journey maps. These data were generated through participatory design sessions with workshop participants. Due to the sensitive and potentially identifiable nature of these data, as well as ethical considerations related to participant privacy and consent, the raw data will not be publicly shared. Aggregated findings and illustrative excerpts are presented within the paper to support transparency and reproducibility. Aggregated findings can be made available by the corresponding author upon reasonable request.

## Ethics Statement

This study is based on qualitative anonymous data collected during the workshop *“Inclusive Medical AI: Exploring Diverse Perspectives”*. The workshop involved participatory design sessions with researchers and patient experts.

Ethical review and approval were not required for this study in accordance with the local legislation and institutional requirements. All participants provided informed consent to participate voluntarily and to have anonymised insights used for scientific dissemination. The study followed the ethical principles of the Declaration of Helsinki.

## Licenses and Copyright

S2 Fig. Adapted from: Wang H, Blok V. Why putting artificial intelligence ethics into practice is not enough: Towards a multi-level framework. Big Data & Society. 2025;12:20539517251340620. https://doi.org/10.1177/20539517251340620. This article is distributed under the terms of the Creative Commons Attribution 4.0 License (https://creativecommons.org/licenses/by/4.0/), which permits any use, reproduction, and distribution of the work without further permission, provided the original work is properly attributed as specified on the SAGE Open Access page (https://us.sagepub.com/en-us/nam/open-access-at-sage).

## References

1. Ethics and Governance of Artificial Intelligence for Health: WHO Guidance. 1st ed. Geneva: World Health Organization; 2021.

2. Lekadir K, Frangi AF, Porras AR, Glocker B, Cintas C, Langlotz CP, et al. FUTURE-AI: international consensus guideline for trustworthy and deployable artificial intelligence in healthcare. BMJ. 2025; e081554. doi:10.1136/bmj-2024-081554

3. Maheshwari K, Jedan C, Christiaans I, van Gijn M, Maeckelberghe E, Plantinga M. AI-Inclusivity in Healthcare: Motivating an Institutional Epistemic Trust Perspective. Camb Q Healthc Ethics. 2024. doi:10.1017/S0963180124000215

4. Alowais SA, Alghamdi SS, Alsuhebany N, Alqahtani T, Alshaya AI, Almohareb SN, et al. Revolutionizing healthcare: the role of artificial intelligence in clinical practice. BMC Med Educ. 2023;23: 1–15. doi:10.1186/s12909-023-04698-z

5. Bajwa J, Munir U, Nori A, Williams B. Artificial intelligence in healthcare: transforming the practice of medicine. Future Healthc J. 2021;8: e188–e194. doi:10.7861/fhj.2021-0095

6. Luna F. Identifying and evaluating layers of vulnerability – a way forward. Dev World Bioeth. 2019;19: 86–95. doi:10.1111/dewb.12206

7. Chan SCC, Neves AL, Majeed A, Faisal A. Bridging the equity gap towards inclusive artificial intelligence in healthcare diagnostics. BMJ. 2024;384: q490. doi:10.1136/bmj.q490

8. Chen RJ, Wang JJ, Williamson DFK, Chen TY, Lipkova J, Lu MY, et al. Algorithmic fairness in artificial intelligence for medicine and healthcare. Nat Biomed Eng. 2023;7: 719–742. doi:10.1038/s41551-023-01056-8

9. Chin MH, Afsar-Manesh N, Bierman AS, Chang C, Colón-Rodríguez CJ, Dullabh P, et al. Guiding Principles to Address the Impact of Algorithm Bias on Racial and Ethnic Disparities in Health and Health Care. JAMA Netw Open. 2023;6: e2345050. doi:10.1001/jamanetworkopen.2023.45050

10. Paccoud I, Leist AK, Schwaninger I, van Kessel R, Klucken J. Socio-ethical challenges and opportunities for advancing diversity, equity, and inclusion in digital medicine. Digit Health. 2024;10: 20552076241277705. doi:10.1177/20552076241277705

11. High-Level Expert Group on AI. Ethics Guidelines for Trustworthy AI. European Commission; 2019. Available: https://digital-strategy.ec.europa.eu/en/library/ethics-guidelines-trustworthy-ai

12. Marko JGO, Neagu CD, Anand PB. Examining inclusivity: the use of AI and diverse populations in health and social care: a systematic review. BMC Med Inform Decis Mak. 2025;25: 57. doi:10.1186/s12911-025-02884-1

13. Sikstrom L, Maslej MM, Hui K, Findlay Z, Buchman DZ, Hill SL. Conceptualising fairness: three pillars for medical algorithms and health equity. BMJ Health Care Inform. 2022;29: e100459. doi:10.1136/bmjhci-2021-100459

14. Balagopalan A, Zhang H, Hamidieh K, Hartvigsen T, Rudzicz F, Ghassemi M. The Road to Explainability is Paved with Bias: Measuring the Fairness of Explanations. 2022 ACM Conference on Fairness, Accountability, and Transparency. Seoul Republic of Korea: ACM; 2022. pp. 1194–1206. doi:10.1145/3531146.3533179

15. Challen R, Denny J, Pitt M, Gompels L, Edwards T, Tsaneva-Atanasova K. Artificial intelligence, bias and clinical safety. BMJ Qual Saf. 2019;28: 231–237. doi:10.1136/bmjqs-2018-008370

16. Chen F, Wang L, Hong J, Jiang J, Zhou L. Unmasking bias in artificial intelligence: a systematic review of bias detection and mitigation strategies in electronic health record-based models. J Am Med Inform Assoc. 2024;31: 1172–1183. doi:10.1093/jamia/ocae060

17. Fehr J, Citro B, Malpani R, Lippert C, Madai VI. A trustworthy AI reality-check: the lack of transparency of artificial intelligence products in healthcare. Front Digit Health. 2024;6. doi:10.3389/fdgth.2024.1267290

18. Goisauf M, Cano Abadía M, Akyüz K, Bobowicz M, Buyx A, Colussi I, et al. Trust, Trustworthiness, and the Future of Medical AI: Outcomes of an Interdisciplinary Expert Workshop. J Med Internet Res. 2025;27: e71236. doi:10.2196/71236

19. Cobanaj M, Corti C, Dee EC, McCullum L, Boldrini L, Schlam I, et al. Advancing equitable and personalized cancer care: Novel applications and priorities of artificial intelligence for fairness and inclusivity in the patient care workflow. Eur J Cancer. 2024;198: 113504. doi:10.1016/j.ejca.2023.113504

20. Decker MC, Wegner L, Leicht-Scholten C. Procedural fairness in algorithmic decision-making: the role of public engagement. Ethics Inf Technol. 2024;27: 1. doi:10.1007/s10676-024-09811-4

21. Griffin AC, Wang KH, Leung TI, Facelli JC. Recommendations to promote fairness and inclusion in biomedical AI research and clinical use. J Biomed Inform. 2024;157: 104693. doi:10.1016/j.jbi.2024.104693

22. Baumgartner R, Arora P, Bath C, Burljaev D, Ciereszko K, Custers B, et al. Fair and equitable AI in biomedical research and healthcare: Social science perspectives. Artif Intell Med. 2023;144: 102658. doi:10.1016/j.artmed.2023.102658

23. Mittermaier M, Raza MM, Kvedar JC. Bias in AI-based models for medical applications: challenges and mitigation strategies. Npj Digit Med. 2023;6: 1–3. doi:10.1038/s41746-023-00858-z

24. Nazer LH, Zatarah R, Waldrip S, Ke JXC, Moukheiber M, Khanna AK, et al. Bias in artificial intelligence algorithms and recommendations for mitigation. Kalla M, editor. PLOS Digit Health. 2023;2: e0000278. doi:10.1371/journal.pdig.0000278

25. Wang H, Blok V. Why putting artificial intelligence ethics into practice is not enough: Towards a multi-level framework. Big Data Soc. 2025;12: 20539517251340620. doi:10.1177/20539517251340620

26. Chen IY, Pierson E, Rose S, Joshi S, Ferryman K, Ghassemi M. Ethical Machine Learning in Healthcare. Annu Rev Biomed Data Sci. 2021;4: 123–144. doi:10.1146/annurev-biodatasci-092820-114757

27. Gorelik AJ, Li M, Hahne J, Wang J, Ren Y, Yang L, et al. Ethics of AI in healthcare: a scoping review demonstrating applicability of a foundational framework. Front Digit Health. 2025;7. doi:10.3389/fdgth.2025.1662642

28. Shams RA, Zowghi D, Bano M. AI and the quest for diversity and inclusion: a systematic literature review. AI Ethics. 2023 [cited 16 Dec 2024]. doi:10.1007/s43681-023-00362-w

29. Nong P, Hamasha R, Platt J. Equity and AI Governance at Academic Medical Centers. Am J Manag Care. 2024;30: SP468–SP472. doi:10.37765/ajmc.2024.89555

30. Zowghi D, da Rimini F. Diversity and Inclusion in Artificial Intelligence. arXiv; 2023. doi:10.48550/ARXIV.2305.12728

31. Fosch-Villaronga E, Poulsen A. Diversity and Inclusion in Artificial Intelligence. In: Custers B, Fosch-Villaronga E, editors. Law and Artificial Intelligence: Regulating AI and Applying AI in Legal Practice. The Hague: T.M.C. Asser Press; 2022. pp. 109–134. doi:10.1007/978-94-6265-523-2_6

32. Sadeghi Z, Alizadehsani R, Cifci MA, Kausar S, Rehman R, Mahanta P, et al. A review of Explainable Artificial Intelligence in healthcare. Comput Electr Eng. 2024;118: 109370. doi:10.1016/j.compeleceng.2024.109370

33. Rudd J, Igbrude C. A global perspective on data powering responsible AI solutions in health applications. Ai Ethics. 2023; 1–11. doi:10.1007/s43681-023-00302-8

34. Ramzy LM, Monson SP, Chao HW-I, Hileman B, Podewils LJ, Pereira RI. Power Dynamics Perpetuate DEI Inaction: A Qualitative Study of Community Health Clinic Teams. Ann Fam Med. 2024;22: 203–207. doi:10.1370/afm.3099

35. Schroeder D, Chatfield K, Chennells R, Partington H, Kimani J, Thomson G, et al. The Exclusion of Vulnerable Populations from Research. In: Schroeder D, Chatfield K, Chennells R, Partington H, Kimani J, Thomson G, et al., editors. Vulnerability Revisited: Leaving No One Behind in Research. Cham: Springer Nature Switzerland; 2024. pp. 25–47. doi:10.1007/978-3-031-57896-0_2

36. Framework for Innovation - Design Council. [cited 28 Mar 2025]. Available: https://www.designcouncil.org.uk/our-resources/framework-for-innovation/

37. Systemic Design Framework - Design Council. [cited 28 Mar 2025]. Available: https://www.designcouncil.org.uk/our-resources/systemic-design-framework/

38. Design Council, The Point People. System-shifting design - An emerging practice explored. UK; 2021 Oct. Available: https://www.designcouncil.org.uk/fileadmin/uploads/dc/Documents/Systemic%20Design%20Report.pdf

39. Roberts JP, Fisher TR, Trowbridge MJ, Bent C. A design thinking framework for healthcare management and innovation. Healthcare. 2016;4: 11–14. doi:10.1016/j.hjdsi.2015.12.002

40. Ryan M, de Roo N, Wang H, Blok V, Atik C. AI through the looking glass: an empirical study of structural social and ethical challenges in AI. AI Soc. 2024 [cited 16 Dec 2024]. doi:10.1007/s00146-024-02146-0

41. Small C, Bjorkegren M, Erkkilä T, Shaw L, Megill C. Polis: Escalar de la deliberación mediante el mapeo de espacios de opinión de alta dimensión. Recer Rev Pensam Anàlisi. 2021;26. doi:10.6035/recerca.5516

42. Miro visual workspace. Miro; 2025. Available: www.miro.com

43. Dagan N, Devons-Sberro S, Paz Z, Zoller L, Sommer A, Shaham G, et al. Evaluation of AI Solutions in Health Care Organizations — The OPTICA Tool. NEJM AI. 2024;1: AIcs2300269. doi:10.1056/AIcs2300269

44. Zuchowski LC, Zuchowski ML, Nagel E. A trust based framework for the envelopment of medical AI. Npj Digit Med. 2024;7: 1–11. doi:10.1038/s41746-024-01224-3

45. Reddy S, Rogers W, Makinen V-P, Coiera E, Brown P, Wenzel M, et al. Evaluation framework to guide implementation of AI systems into healthcare settings. BMJ Health Care Inform. 2021;28: e100444. doi:10.1136/bmjhci-2021-100444

46. Goirand M, Austin E, Clay-Williams R. Implementing Ethics in Healthcare AI-Based Applications: A Scoping Review. Sci Eng Ethics. 2021;27: 1–53. doi:10.1007/s11948-021-00336-3

47. Delgado F, Yang S, Madaio M, Yang Q. The Participatory Turn in AI Design: Theoretical Foundations and the Current State of Practice. Proceedings of the 3rd ACM Conference on Equity and Access in Algorithms, Mechanisms, and Optimization. New York, NY, USA: Association for Computing Machinery; 2023. pp. 1–23. doi:10.1145/3617694.3623261

48. The Algorithm Register of the Dutch government. [cited 15 Sept 2025]. Available: https://algoritmes.overheid.nl/en

49. Article 4: AI literacy | EU Artificial Intelligence Act. [cited 15 Sept 2025]. Available: https://artificialintelligenceact.eu/article/4/

50. Samuel G, Ballard LM, Carley H, Lucassen AM. Ethical preparedness in health research and care: the role of behavioural approaches. BMC Med Ethics. 2022;23: 115. doi:10.1186/s12910-022-00853-1

51. Wang H, Blok V, van Hilten M. ELSA Labs for responsible AI: a novel approach for addressing ethical, legal, social issues. J Responsible Innov. 2025;12: 2563944. doi:10.1080/23299460.2025.2563944

